# Risk factors associated with severe outcomes of COVID-19: A systematic rapid review to inform national guidance on vaccine prioritization in Canada

**DOI:** 10.1101/2021.04.23.21256014

**Authors:** Michelle Gates, Jennifer Pillay, Aireen Wingert, Samantha Guitard, Sholeh Rahman, Bernadette Zakher, Allison Gates, Lisa Hartling

## Abstract

**Background:** To inform vaccine prioritization guidance in Canada, we systematically reviewed evidence on the magnitude of association between risk factors and severe outcomes of COVID-19. The urgent nature of this review necessitated an adapted methodology, which may serve as an exemplar for reviews undertaken under strict timelines.

**Methods:** We updated our existing review by searching online databases and websites for cohort studies providing multivariate adjusted associations. After piloting, one author screened studies and extracted data. Two authors estimated the magnitude of association between exposures and outcomes as little-to-no (odds, risk, or hazard ratio <2.0, or >0.50 for reduction), large (2.0-3.9, or 0.50-0.26 for reduction), or very large (≥4.0, or ≤0.25 for reduction), and rated the evidence certainty using GRADE.

**Results:** Of 11,734 unique records we included 134 reports. There is probably (moderate certainty) at least a large increase in mortality from COVID-19 among people aged 60-69 vs. <60 years (11 studies, n=517,217), with ≥2 vs. no comorbidities (4 studies, n=189,608), and for people with (vs. without): Down syndrome (1 study, n>8 million), type 1 and 2 diabetes (1 study, n>8 million), end-stage kidney disease (1 study, n>8 million), motor neuron disease, multiple sclerosis, myasthenia gravis, or Huntington’s disease (as a grouping; 1 study, n>8 million). The magnitude of association with mortality is probably very large for Down syndrome and may (low certainty) be very large for age 60-69 years, and diabetes. There is probably little-to-no increase in severe outcomes with several cardiovascular and respiratory conditions, and for adult males vs. females.

**Conclusion:** There is strong evidence to support at least a large increase in mortality from COVID-19 among older adults aged 60 to 69 years versus <60 years; people having two or more versus no comorbidities; and for people affected by several pre-existing conditions. The methodology employed in this review may provide an important exemplar for future syntheses undertaken under urgent timelines.

**Systematic review registration:** PROSPERO #CRD42021230185.

## INTRODUCTION

The novel coronavirus disease 2019 (COVID-19) became a worldwide public health concern in early 2020 [1]. About 20% of affected people experience severe disease (e.g., requiring hospitalization) [2], and some may have long-lasting symptoms or medical complications [3]. In late 2020, Health Canada approved two vaccines for the prevention of COVID-19 (Pfizer-BioNTech COVID-19 Vaccine and Moderna COVID-19 Vaccine); this was been followed by the approval of others and continued review of emerging vaccine candidates [4]. As anticipated, vaccine distribution in Canada was initially constrained by logistical challenges and limited supply [5–7], necessitating prioritization of populations for vaccination [6, 7].

The National Advisory Committee on Immunization (NACI) developed preliminary vaccine prioritization guidance in late 2020 “*for the efficient, effective and equitable allocation of safe, efficacious severe acute respiratory syndrome coronavirus 2 (SARS-CoV-2) vaccines in the context of staggered arrival of vaccines”* [6, 7]. In addition to reviewing evidence on the burden of illness and vaccine characteristics, NACI used its Ethics, Equity, Feasibility and Acceptability (EEFA) framework [8] and evidence-informed Equity Matrix [9] to achieve the goal of their prioritization guidance. The Equity Matrix considers biological and social risk factors (termed P^2^ROGRESS And Other Factors, informed by the PROGRESS-Plus model [10]) that may result in inequitable health outcomes across a population. The findings of a rapid review of risk factors for severe outcomes of COVID-19 conducted by our research group [11] helped populate the Equity Matrix, and informed NACI’s guidance. This initial review found increasing age (>60 years and particularly >70 years) to be among the strongest risk factors for severe outcomes of COVID-19 [11].

By early 2021, numerous additional relevant primary studies had been published. To inform subsequent guidance statements [12, 13], we updated our rapid review to determine the magnitude of association between P^2^ROGRESS And Other Factors and severe outcomes of COVID-19. Though vaccines for COVID-19 are now widely available to Canadians, this review serves an important archival function in informing vaccine prioritization in regions where distribution is less far along. The methodology used in this review may serve as an exemplar for similar high-priority scenarios where syntheses of large volumes of data from primary studies are needed on expedited timelines.

## METHODS

### Review Approach

Detailed methods are in our pre-defined protocol (PROSPERO #CRD42021230185), and described briefly herein. We followed traditional systematic review methods [14] with modifications to accommodate the expedited timeline needed to inform NACI’s guidance. Single experienced reviewers selected studies and extracted data, with piloting to maintain rigour. We restricted the scope to large (>1,000 participants) studies with a high relevance to Canada.

The eligibility criteria were informed by stakeholders at NACI and the Public Health Agency of Canada; these individuals were not involved in review conduct. This review is reported according to the Preferred Reporting Items for Systematic Reviews and Meta-Analyses (PRISMA) statement [15].

### Eligibility criteria

Table 1 details our eligibility criteria. We included published (or accepted for publication) cohort studies reporting multivariate adjusted associations between P^2^ROGRESS And Other Factors (including but not limited to age; gender identity or sex; disease/condition or disability; number of comorbidities; social factors; other factors such as health behaviours) and severe outcomes of COVID-19 among: the general population; people with COVID-19; people hospitalized with COVID-19; people with severe COVID-19 (as defined by study authors; typically in an intensive care unit and/or mechanically ventilated); children in any of the aforementioned populations. Outcomes of interest were: hospitalization; in-hospital length of stay; ICU admission; ICU length of stay; mechanical ventilation; mortality (all-cause or case fatality); severe disease (composite outcomes as defined by study authors, most often combining mortality with ICU admission and/or mechanical ventilation); need for rehabilitation; stroke; kidney, liver, or cardiac injury; generic functionality and/or disability (composite scores of validated scales); generic quality of life (composite scores of validated scales). For age, we included data from all relevant comparisons, however were interested specifically in the comparison of age 60-69 years vs. <60 years, as our previous review already showed older age (particularly >70 years) to be among the strongest risk factors for severe outcomes of COVID-19 [11]. Canadian reports of any design with any analysis type were eligible. For social risk factors, we included only Canadian reports because these were considered as most relevant by NACI.

**Table 1.** Eligibility criteria for inclusion of studies in the review

|  | **Inclusion and exclusion criteria** |
| --- | --- |
| **Study design** | **Include:**<br>– Prospective and retrospective cohort studies, including registry-based studies (may be published as a letter); government reports; pre-prints that have been accepted for publication<br>– Canadian reports of any design<br>– Systematic reviews, only for risk factors of high relevance to NACI where no primary studies are found<br>**Exclude:** Pre-prints that have not been accepted by a peer-reviewed journal, commentaries, editorials |
| **Sample size** | Primary studies were required to include at least 1,000 participants (≥300 for studies of children). There was no sample size requirement for systematic reviews. |
| **Population** | **Include:** People of any age in the general population; with COVID-19 infection (laboratory-confirmed or epidemiologically linked); hospitalized with COVID-19; with severe COVID-19 (as defined by study authors); children in any of the aforementioned populations.<br>**Exclude:** Populations with other pandemic-related infections, if data for COVID-19 cannot be isolated. People with COVID-19 that is not laboratory-confirmed or epidemiologically linked. When unclear, we included studies if it appeared that the large majority (ie., ≥90%) would be confirmed cases. |
| **Exposure** | **Include:** Any P<sup>2</sup>ROGRESS And Other Factors including but not limited to age; gender identity or sex; disease/condition or disability; number of comorbidities; social factors (place/state of residence; race, ethnicity, immigrant/refugee status, Indigenous identity, culture, language; occupation; religion, belief system; education, literacy; socioeconomic status; social capital); other (e.g., health behaviours).<br>**Exclude:** Exposures that constitute laboratory findings, vital signs, or symptoms occurring after COVID-19 infection; any other exposures not of interest to NACI. |
| **Comparator** | **Include:** (a) no exposure to the P<sup>2</sup>ROGRESS And Other Factors; (b) the same P<sup>2</sup>ROGRESS And Other Factors experienced differently or to a different extent (e.g., different age group). When included, systematic reviews did not require a comparator.<br>**Exclude:** Variations in disease severity within the pre-existing disease/condition or disability category (i.e., only interested in presence/absence of this risk factor); studies without a comparator. |
| **Outcome** | **Include:** Epidemiologic or analytic data from Canadian reports; from all other studies, magnitude of association (based on multivariate analysis adjusting for at minimum age and sex)* between the exposure and any of hospitalization; in-hospital length of stay; ICU admission; ICU length of stay; mechanical ventilation; mortality (all-cause or case fatality); severe disease (composite outcomes as defined by study authors); need for rehabilitation; stroke; kidney, liver, or cardiac injury; generic functionality and/or disability (composite scores of validated scales); generic quality of life (composite scores of validated scales).<br>*Age and sex needed to be adjusted for at least one pre-existing condition. Pre-existing conditions (except immunocompromised, autoimmune, neurologic, mental health, frailty) needed to be adjusted for body mass index. <sup>b</sup><br>**Exclude:** All other outcomes; disease-specific functionality, disability, or quality of life; crude / unadjusted associations; studies where the focus is symptoms, biomarkers, clinical variables collected after disease onset. |
| **Timing** | Any length of follow-up; at least 28 days for mortality |
| **Setting** | – For social risk factors among adults, only Canadian studies<br>– For all other risk factors, Organization for Economic Cooperation and Development countries: https://www.oecd.org/about/document/list-oecd-member-countries.htm<br>– Systematic reviews could include studies from any country |
| **Language** | English or French |

COVID-19=novel coronavirus 2019; ICU=intensive care unit; NACI=National Advisory Committee on Immunization
<sup>a</sup> The main scope differences versus the original review were: addition of a sample size requirement; inclusion of Canadian reports of any design; addition of children as an important population; inclusion of Canadian reports only for social factors; requirement of adjustment for more than age and sex for some risk factors; addition of organ injury and long-term outcomes of relevance to NACI.
<sup>b</sup> Decisions for adjustment by body mass index were informed by the findings of our previous review and expert input from the Public Health Agency of Canada.

We applied criteria at the selection stage to include only the most informative studies. Studies needed to take place in Organization for Economic Cooperation and Development (OECD) countries and include ≥1,000 participants (i.e., suitable for multivariate adjustment). Adjustment by age and sex was required in all studies, except Canadian reports. Findings for age and/or sex required adjustment for at least one pre-existing condition, and findings for most pre-existing conditions required adjustment for body mass index (BMI). After screening, we loosened our selection criteria for studies among children, requiring a sample size of ≥300 participants and adjustment by age and sex (but not BMI), as limited evidence was available for this population.

When no primary studies were located on a risk factor of high relevance to NACI, we included systematic reviews with broader selection criteria.

### Literature search and study selection

Full details of the literature search are in Additional file 1. A research librarian searched online databases (Ovid Medline® ALL 1946-, Epistemonikos COVID-19 in L-OVE Platform [https://app.iloveevidence.com/loves] for individual predictors of outcomes; 2-3 December 2020) and websites suggested by NACI (6 January - 3 February, 2021). We updated the Medline and website searches in April 2021 for the pediatric population, immune compromise, and autoimmune conditions, to further inform NACI’s guidance for risk factors. We exported records to Endnote X9 (Clarivate Analytics, Philadelphia, PA), removed duplicates and pre-prints, then uploaded the records to Covidence systematic review platform (https://www.covidence.org/). After piloting (i.e., practice round with all team members), a single experienced reviewer screened each record by title and abstract, then by full text. A second reviewer was consulted if assistance was required to determine whether a study met the selection criteria.

Following screening, gaps in evidence (i.e., no study meeting our criteria) were apparent for asplenia/splenic dysfunction, cystic fibrosis, sickle cell disease, β-thalassemia, and learning disability (potentially important based on inclusion in priority lists from other jurisdictions [16–19]). To locate evidence for these, we scanned all studies excluded based on sample size (<1,000 participants), preprints, and systematic reviews that we had located during our scoping exercises (September, 2020) using Smart Searches in Endnote. On 18 February 2021, we conducted targeted searches in Ovid Medline® ALL 1946-for studies and reviews on these risk factors.

### Data extraction

After piloting (i.e., practice round with all team members), a single reviewer extracted data from each study into Excel (v. 2016; Microsoft Corporation, Redmond, WA): study and population characteristics, exposures, covariates, outcomes and their definitions, findings for multivariate associations. We held regular team meetings to troubleshoot and ensure consistency. A second reviewer was consulted to review and confirm ambiguous data presented within the included studies.

### Quality assessment

Because we attempted to include only good quality studies (i.e., large sample size and with adequate multivariate adjustment), we did not assess their quality using a formal tool. Instead, we recorded potential risk of bias concerns (Additional file 2) during data extraction, and considered these during our assessment of the certainty of evidence. The items (potential concerns) recorded included but were not limited to: (a) lack of adjustment for social factors; (b) the expectation of attenuated magnitude of association when the model was adjusted for laboratory values and/or symptoms; (c) for the hospitalization outcome, potential for testing bias in certain populations (e.g., healthcare providers), where the exposure group may have had less severe disease than those not exposed; (d) use of historical, potentially inaccurate, data for risk factors or covariates.

### Synthesis and drawing conclusions

Additional file 2 details our approach to synthesis and drawing conclusions. Results from the Canadian studies, none of which met the criteria of having sufficiently adjusted analysis, were not included in the main synthesis and are reported separately. We did not pool findings statistically for any outcome, due to large heterogeneity in comparisons and measures of association, and at least some overlap in populations across studies. Instead, two reviewers reached consensus on a best estimate of the magnitude of association for each outcome-comparison across the relevant studies (based on adjusted odds [aOR], risk, or hazard ratios): little-to-no (<2.0, or >0.5 for reduction), large (2.0-3.9, or 0.5-0.26 for reduction), or very large (≥4.0, or ≤0.25 for reduction). Two reviewers assessed the certainty of evidence for each exposure-outcome association, informed by elements of the Grading of Recommendations, Assessment, Development and Evaluation approach [20], and agreed on the final rating. Given our focus on prognostic variables, findings from observational studies for each outcome-comparison started at high certainty [21], and were rated down for concerns related to risk of bias, inconsistency, imprecision, and indirectness [20].

## RESULTS

### Included studies

Of 11,734 unique records identified, we included 134 reports (Figure 1) [22–155]. Full text exclusions are listed at [https://doi.org/10.7939/DVN/W1M6OP]. We included one systematic review of case series for β-thalassemia [142]. No studies or reviews were located with relevant findings for asplenia/splenic dysfunction, cystic fibrosis, sickle cell disease, or learning disability. No study reported on long-term outcomes. Table 2 shows a summary of the characteristics of the studies presenting multivariate adjusted findings that were included in the main synthesis (n=123) [22–144]; full details by study, including 11 Canadian reports [145–155], are in Additional file 3. The studies contributing multivariate associations originated primarily from the United States (n=55), the United Kingdom (n=15), and Mexico (n=12), and included more than 76 million participants (median 5,279; range 34 to 61,414,470).

**Figure 1.**
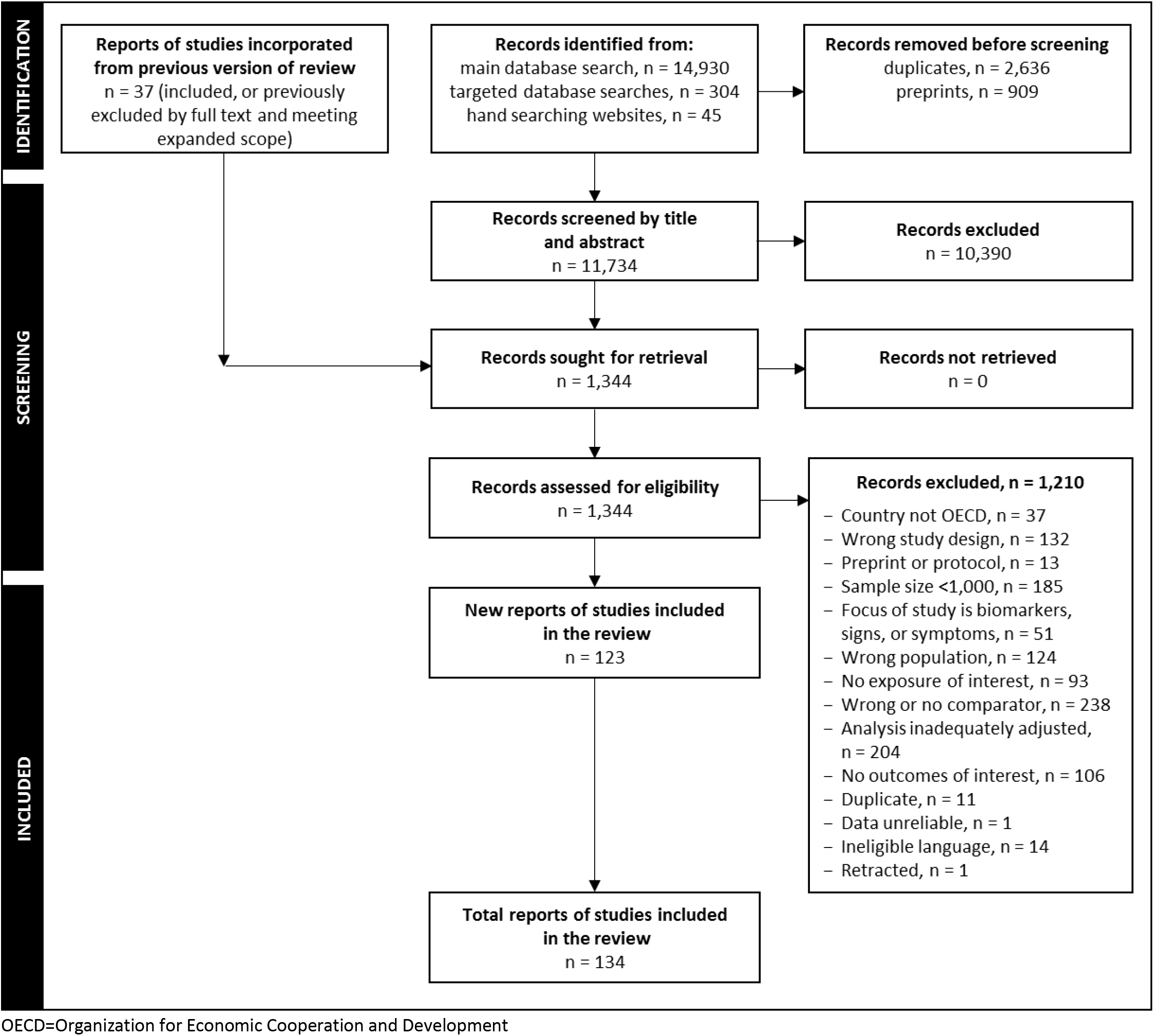
Flow of records through the selection process

**Table 2.** Summary of study characteristics for the studies contributing to the main synthesis, n=123

| Characteristics | Number of studies | Proportion of studies (%) |
| --- | --- | --- |
| STUDIES |  |  |
| Design |  |  |
| Retrospective cohort | 92 | 74.8 |
| Prospective cohort | 29 | 23.6 |
| Nested case-control | 1 | 0.8 |
| Systematic review <sup>b</sup> | 1 | 0.8 |
| Setting |  |  |
| United States | 55 | 44.7 |
| United Kingdom | 15 | 12.2 |
| Mexico | 12 | 9.8 |
| Italy | 9 | 7.3 |
| Korea | 6 | 4.9 |
| Spain | 6 | 4.9 |
| Denmark | 4 | 3.3 |
| France | 3 | 2.4 |
| Other <sup>c</sup> | 16 | 13.0 |
| Funding source |  |  |
| Non-industry | 58 | 47.2 |
| Industry | 3 | 2.4 |
| No funding | 28 | 22.8 |
| Not reported | 36 | 29.3 |
| PARTICIPANTS <sup>d</sup> |  |  |
| Number of participants, median (range) | 5,279 (34 to 61,414,470) |  |
| Mean or median age in years, median (range) | 62.0 (5.0 to 78.6) |  |
| Sex/gender, median % male (range) | 49.8 (0 to 94.0) |  |
| Population analyzed |  |  |
| General population | 10 | 8.1 |
| People with COVID-19 | 59 | 48.0 |
| Hospitalized with COVID-19 | 62 | 50.4 |
| With severe disease (in ICU or mechanically ventilated) | 9 | 7.3 |
| COVID-19 ascertainment |  |  |
| RT-PCR | 94 | 76.4 |
| Laboratory confirmed (unspecified test) | 25 | 20.3 |
| Antigen test | 3 | 2.4 |
| Nucleic acid test | 2 | 1.6 |
| Rapid antibody test | 2 | 1.6 |
| EXPOSURES |  |  |
| Pre-existing conditions | 95 | 77.2 |
| Gender identity/sex | 70 | 56.9 |
| Age (60-69 versus <60 years) | 41 | 33.3 |
| Other (health behaviours) | 27 | 22.0 |
| Number of comorbidities | 21 | 17.1 |
| Children | 11 | 8.9 |
| Exposure - occupation | 4 | 3.3 |
| Pregnancy | 4 | 3.3 |
| OUTCOMES |  |  |
| Hospitalization | 36 | 29.3 |
| In-hospital length of stay | 4 | 3.3 |
| ICU admission | 27 | 22.0 |
| ICU length of stay | 1 | 0.8 |
| Mechanical ventilation | 22 | 17.9 |
| Acute kidney injury | 8 | 6.5 |
| Ischemic stroke | 1 | 0.8 |
| Severe disease | 28 | 22.8 |
| Mortality | 88 | 71.6 |

COVID-19=novel coronavirus disease 2019; ICU=intensive care unit; RT-PCR=reverse transcription polymerase chain reaction
<sup>a</sup> Categories do not all add to 100%, as it is possible for studies to report on more than one category (e.g., used more than one COVID-19 ascertainment method, analyzed more than one population).
<sup>b</sup> Included for β-thalassemia as no primary studies meeting the selection criteria were located.
<sup>c</sup> Includes Belgium (n=1), , Germany (n=2), Ireland (n=1), multi-country (n=3), Netherlands (n=1), Sweden (n=2), Turkey (n=2), UK and Italy (n=1).
<sup>d</sup> 24 studies did not report on patient mean or median age, one study in children did not report on patient sex/gender.

### Associations between risk factors and outcomes

Based on stakeholder input, we considered people with confirmed COVID-19 to be the population of highest relevance, thus have focused on these in our presentation of findings. When no data were available from the COVID-19 population for a risk factor-outcome comparison, we relied on findings from the general population. Table 3 summarizes the risk factors showing large or very large multivariate-adjusted associations with severe outcomes. Additional file 2 includes findings for all risk factors and populations investigated in the review. Data supporting the conclusions, including precise associations presented in each included study, are available at [https://doi.org/10.7939/DVN/W1M6OP].

**Table 3.** Risk factors identified that have large or very large associations with severe outcomes of COVID-19 and the corresponding level of certainty in the association

| Risk factor (among people with COVID-19 unless otherwise specified) | Outcome of interest | Magnitude of association <sup>a</sup><br>(certainty in association) <sup>b</sup> |
| --- | --- | --- |
| **Age** |  |  |
| 60-69 vs. <60 years | Hospitalization | Large (moderate) |
|  | Mechanical ventilation | Large (low) |
|  | Severe disease | Large to very large (low) |
|  | Mortality | Large/very large<br>(moderate/low) |
| **Pre-existing conditions** |  |  |
| 2 or more vs. no comorbidities | Hospitalization | Large (moderate) |
|  | ICU admission | Large (low) |
|  | Mechanical ventilation | Large (low) |
|  | Mortality | Large (moderate) |
| Down syndrome <sup>c</sup> | Hospitalization | Large/very large<br>(moderate/low) |
|  | Mortality | Very large (moderate) |
| Type 1 diabetes <sup>c</sup> | Mortality | Large/very large<br>(moderate/low) |
| Type 2 diabetes <sup>c</sup> | Mortality | Large/very large<br>(moderate/low) |
| End-stage kidney disease <sup>c</sup> | Mortality | Large (moderate) |
| Motor neuron disease, multiple sclerosis, myasthenia gravis, or Huntington's disease <sup>c, d</sup> | Mortality | Large (moderate) |
| Chemotherapy in the past 12 months (Grade A-C) <sup>c, e</sup> | Mortality | Large/very large (low) |
| Radiotherapy in the past 6 months <sup>c</sup> | Mortality | Large (low) |
| Among people <70 years, metastatic cancer | Mortality | Large (low) |
| Parkinson's disease <sup>c</sup> | Mortality | Large (low) |
| Sickle cell disease or severe immunodeficiency <sup>c</sup> | Mortality | Large (low) |
| Solid organ transplant | Mortality | Large (low) |
| Transplant (any, including solid organ) | Hospitalization | Large (low) |
| Kidney transplant (due to stage 5 kidney disease) <sup>c</sup> | Mortality | Large (low) |
| Recent bone marrow or stem cell transplant <sup>c</sup> | Mortality | Large (low) |
| Vasculitis <sup>c</sup> | Hospitalization | Large (low) |
| Major psychiatric disorder (schizophrenia, schizoaffective disorder, or bipolar disorder) <sup>f</sup> | Mortality | Large (low) |
|  | Hospitalization | Large (low) |
| Cerebral palsy <sup>c</sup> | Mortality | Large (low) |
| Diabetes (any type) in females | Mortality | Large (low) |
| Obesity class III vs. normal weight | Mortality | Large (low) |
| Previous cerebrovascular accident | Hospitalization | Large (moderate) |
| Obesity (all classes) vs. normal weight <sup>c</sup> | Hospitalization | Large (low) |
| Obesity class III vs. lower body mass index | Hospitalization | Large (low) |
| Obesity hypoventilation syndrome | Mechanical ventilation | Large (low) |
| Pregnancy (any stage) | Hospitalization | Large (low) |
|  | ICU admission | Large (low) |
|  | Severe disease (pneumonia) | Large (low) |
| Frailty <sup>c, g</sup> | Hospitalization | Large (low) |
| Chronic kidney disease | Hospitalization | Approaching large (low) |
| Diabetes (any) | Hospitalization | Approaching large (low) |
| **Children (as defined by individual studies)** |  |  |

| **Risk factor (among people with COVID-19 unless otherwise specified)** | **Outcome of interest** | **Magnitude of association <sup>a</sup>**<br>**(certainty in association) <sup>b</sup>** |
| --- | --- | --- |
| Children (0-19 years) vs. adults (50-59 years) | Hospitalization | Large reduction (low) |
| Age <1 month vs. >1 month | ICU admission | Large (low) |
| Immunodeficiency or immunosuppression (unspecified reason) | Mortality | Large to very large (low) |
|  | Hospitalization | Large (low) |
| Hypertension | Hospitalization | Large (low) |
| 2 or more vs. no chronic conditions <sup>h</sup> | Hospitalization | Large (moderate) |
| 1 vs. no chronic conditions <sup>h</sup> | Hospitalization | Large (low) |
| 1 or more vs. no chronic conditions <sup>i</sup> | ICU admission | Large (low) |
|  | Mechanical ventilation | Large (low) |
|  | Severe disease | Large (low) |
| Asthma | ICU admission | Large (low) |
| Endocrine condition <sup>j</sup> | Severe disease | Large (low) |
| Metabolic condition <sup>j</sup> | Severe disease | Large (low) |
| Malignancy <sup>j</sup> | Severe disease | Large (low) |
| Black race/ethnicity vs. White non-Hispanic | Mortality | Large (low) |
|  | Hospitalization | Large (low) |
| **Risk of exposure - occupation** |  |  |
| Healthcare workers (vs. non-healthcare workers) | Hospitalization | Large reduction (low) |
|  | ICU admission | Large reduction (low) |

ICU=intensive care unit; vs.=versus
<sup>a</sup> Magnitude of associations are shown as large (OR or RR $\geq 2.00$ , or $\leq 0.5$ for reduction) or very large (OR or RR $\geq 4.00$ ).
<sup>b</sup> Confidence in the magnitude of the associations was determined by considering primarily consistency in findings across studies, directness of the setting and risk factors (e.g., type of healthcare system, uncertainty about risk factor clearly matching review criteria), precision (e.g., confidence intervals indicating possibility of little to no association), and potential risk of bias. Low confidence indicates that there may be an association and moderate means that the evidence indicates that there probably is an association of the magnitude stated.
<sup>c</sup> The denominator for these risk factors is the general population. These were included when findings for the risk factor-outcome comparison were not available from studies of populations with COVID-19.
<sup>d</sup> These conditions were grouped within a single study; evidence for the individual conditions is either unavailable or of lower certainty.
<sup>e</sup> There was evidence of a large to very large increase in mortality with grades B and C chemotherapy from one large study. However, the stages of chemotherapy were not defined, and could not be ascertained from the study's authors. In the absence of adequate information, we have grouped all stages of chemotherapy (A, B, and C) together for analysis.
<sup>f</sup> Defined by hospital discharge diagnosis, in combination with drug use (filled a prescription) for the condition in the past 6 months.
<sup>g</sup> General population sample that may include community and non-community dwelling people. Measured on scales that include items such as weight loss, exhaustion, physical activity, walking speed, grip strength, overall health, disability, presence of disease, dementia, falls, mental wellbeing.
<sup>h</sup> Includes congenital malformations, asthma, epilepsy, complex genetic syndromes, endocrine disorders, cancers, hematologic diseases, rheumatic diseases, autoimmune disease, autism or neurologic development impairment, gastrointestinal diseases, liver disease, renal disease, genitourinary diseases, cystic fibrosis or other chronic lung diseases, metabolic disorders, hydrocephalus, severe obesity, hypertension, otolaryngologic diseases, pregnancy, and 'complex chronic conditions'.

The certainty of evidence was moderate for at least a large increase in mortality from COVID-19 for people aged 60-69 years versus <60 years (11 studies, n=517,217) [27, 57, 72, 78, 82, 97, 112, 113, 118, 121, 127, 135], people with two or more versus no comorbidities (4 studies, n=189,608) [57, 68, 121, 126], and for people with (versus without): Down syndrome (1 study, n>8 million) [41], type 1 and 2 diabetes (1 study, n>8 million) [41], end-stage kidney disease (1 study, n>8 million) [41]; motor neuron disease, multiple sclerosis, myasthenia gravis, or Huntington’s disease (as a grouping; 1 study, n>8 million) [41]. The magnitude of association with mortality is probably (moderate certainty) very large for Down syndrome, and may (low certainty) be very large for age 60-69 years, and diabetes. We located no evidence on the combination of conditions that would place people with two or more comorbidities at increased risk.

The certainty of evidence was low for a large increase in mortality or hospitalization among: people with cerebral palsy (1 study, n>8 million); major psychiatric disorder (schizophrenia, schizoaffective disorder, or bipolar disorder, with drug use for the condition in the past 6 months; 1 study, n=11,122) [126]; obesity class III (BMI ≥40 kg/m^2^) versus normal weight (1 study, n=1,612) [135]; Parkinson’s disease (1 study, n>8 million) [41]; sickle cell disease or severe immunodeficiency (1 study, n>8 million) [41]; solid organ transplant (2 studies, n=18,038 for mortality [126, 135]); transplant of any type (3 studies, n=24,227 for hospitalization [58, 74, 126]); kidney transplant (1 study, n>8 million) [41]; recent bone marrow or stem cell transplant (1 study, n>8 million) [41]; metastatic cancer among those <70 years (2 studies, n=38,377) [67, 135]; current or recent (past year) chemotherapy or radiotherapy (past 6 months; 1 study, n>8 million); vasculitis (1 study, n=4,543,249) [42]; pregnancy (any stage; 1 study, n=409,462) [40]; frailty (2 studies, n=404,079) [89, 115]. The magnitude of association with mortality may (low certainty) be very large for recent chemotherapy. There was moderate certainty for a large increase in hospitalization with previous cerebrovascular accident (2 studies, n=12,376) [58, 138].

Among children, there was evidence of moderate certainty for a large increase in hospitalization among those having two or more versus no chronic conditions (1 study, n=804) [52], and evidence of low certainty for a large increase in mortality and hospitalization for children of Black versus White non-Hispanic race/ethnicity (1 study, n=12,198) [106]. There was low certainty for a large reduction in hospitalization for children 0-19 years versus adults aged 50-59 years (1 study, n=2,199) [126], and a large increase in hospitalization for children with one versus no chronic conditions (1 study, n=790) [52], and specifically for children with versus without immunodeficiency or immunosuppression (2 studies, n=21,875) [55, 94], and hypertension (1 study, n=21,116) [94]. Additionally, there was evidence of low certainty for a large increase in severe outcomes (ICU admission, mechanical ventilation, or ‘severe disease’) among children with versus without: one or more chronic conditions (1 study, n=759 for mechanical ventilation and severe disease [55]; 2 studies, n=1,341 for ICU admission), asthma (1 study, n=21,116) [94], endocrine conditions (1 study, n=5,374) [32], metabolic conditions (1 study, n=5,374) [32], and malignancy (1 study, n=5,374) [32].

There were several risk factors with moderate certainty evidence for little-to-no association with increased severity of COVID-19, for example many pre-existing conditions (notably several cardiovascular and respiratory conditions), and for adult males versus females (Additional file 2).

Table 4 shows the risk factors identified in Canadian reports and potential associations with severe outcomes of COVID-19. All data collected from these studies are in Additional file 2. This evidence was used primarily to draw conclusions about social factors, and was often descriptive and lacking adjustment for important covariates. These studies showed the potential for a large increase in mortality among people living in long-term care (6 reports, n≈2 million) [145, 146, 148, 151, 153], visible minority groups (mainly South Asian, Chinese, Black, Filipino, Latin American, Arab, Southeast Asian, West Asian, Korean, Japanese; 1 report, n=8,796) [154], and people living in the general population (off-reserve) versus First Nations people living on-reserve (1 report, n=9,715) [147].

**Table 4.** Risk factors identified in Canadian reports and potential associations with severe outcomes of COVID-19

| **Risk factor** | **Reports (participants)** | **Conclusion** |
| --- | --- | --- |
| **Social factors** |  |  |
| Living in long-term care | 6 (≈2 million) | Probably associated with a large increase in mortality; the association may be very large for those in their 60s and 70s. |
| Being part of a visible minority population <sup>a</sup> | 1 (8,796) | May be associated with increased mortality; magnitude uncertain and findings rely on ecological data and did not account for important covariates. |
| Living on a First Nations reserve | 1 (9,715) | Rates of hospitalization and mortality appear lower compared to living off-reserve. The evidence does not account for potentially important covariates such as age. |
| Being a healthcare worker | 2 (23,101) | Among those aged 30-70 years, may be associated with a large reduction in hospitalization, ICU admission, and mortality. Among healthcare workers, males appear to have higher rates of hospitalization, ICU admission, and mortality than females. Analyses did not account for important covariates. |
| Homelessness | 2 (18,224) | Very uncertain about associations with mortality. |
| Being a homeless shelter worker | 1 (1,734) | Very uncertain about associations with mortality. |
| **Age and gender identity/sex** |  |  |
| Age 60-69 vs. <60 years | 5 (235,481) | May be associated with a large increase in hospitalization, ICU admission, and mortality. For mortality there may also be a large association when compared to 50-59 years. The magnitude of association appears to account for pre-existing medical conditions and seems similar between sexes, though absolute rates are high among men in all age groups, particularly among those not residing in long-term care or delivering healthcare. Findings for hospitalization and ICU admission may be most applicable to those living outside of long-term care as there has been limited transfer to hospital and ICU from this setting. |
| Male vs. female | 2 (11,259) | Rates of hospitalization and ICU admission appear to be lower among females >30 years; after excluding health care workers, long-term care residents, and school/daycare workers and attendees, the reduction in hospitalization and ICU admission was only observed in those >50 years. Case fatality appears to be lower among females than males in the same age group. |
| **Pre-existing conditions** |  |  |
| One vs. no comorbidities | 2 (15,875) | In all age groups, may have little-to-no association with mortality. |
| Two or more vs. <2 comorbidities | 1 (6,350) | Among people aged 60-79 years, may be associated with a large increase in mortality. Study data were not adjusted for important covariates like residing in long-term care. Findings for people <60 years were limited by small sample size. |
| Compromised immunity | 1 (1,734) | May be associated to some extent with an increase in mortality; the magnitude of association is uncertain due to a wide confidence interval. This association was adjusted for age, long-term care residency, smoking, renal disease, diabetes, and chronic obstructive pulmonary disease. |
| Malignancy | 2 (11,259) | May be associated to some extent with an increase in mortality; the magnitude of association is uncertain due to lack of adjustment for important covariates. |
| Pre-existing chronic conditions | 2 (11,259) | Chronic obstructive pulmonary disease, diabetes, and renal disease may be associated with a large increase in mortality; these findings lacked adjustment for sex, BMI, or social factors, which may be important confounders. Other pre-existing conditions that are common among COVID-19 deaths include dementia and Alzheimer's disease, pneumonia, hypertension, ischemic heart disease, respiratory failure, renal failure, chronic lower respiratory disease, nervous system disorders, and cancer. These findings are from epidemiologic data and not adjusted for important covariates. |
| **Special populations** |  |  |
| Pregnancy | 1 (137,901) | Among females of childbearing age with COVID-19, pregnancy may be associated with a large-to-very large increase in hospitalization and ICU admission. The findings were not adjusted for important covariates. |
| **Children and young adults** |  |  |
| Pre-existing disease in young people | 1 (9,525) | Among young adults, adolescents, and children, mortality from COVID-19 may be associated with having at least one comorbidity. This finding is not adjusted for important covariates. |
| Pre-existing disease in infants <1 year old | 1 (27) | Among a small sample of infants with COVID-19, few (20%) who were hospitalized had a comorbid condition. The findings were not adjusted for any covariates. |
| Age in infants <1 year old | 1 (27) | There appeared to be little-to-no difference in severe outcomes by age. The findings were not adjusted for any covariates. |
| Sex/gender in infants <1 year old | 1 (27) | Among a small sample of infants with COVID-19, half of those hospitalized were male and half were female. The findings were not adjusted for any covariates. |

COVID-19=novel coronavirus disease 2019; BMI=body mass index; ICU=intensive care unit
<sup>a</sup> Defined as "Persons, other than Aboriginal peoples, who are non-Caucasian in race or non-white in colour." Includes mainly South Asian, Chinese, Black, Filipino, Latin American, Arab, Southeast Asian, West Asian, Korean, Japanese.

## DISCUSSION

This rapid systematic review provides a methodologically rigorous synthesis of a large volume of emerging evidence on factors associated with severe outcomes of COVID-19. We identified strong (moderate certainty) evidence of at least a large increase in mortality from COVID-19 among people aged 60 to 69 versus <60 years (and over 70 years from our previous review [140]), people having two or more versus no comorbidities, and for people affected by (versus unaffected): Down syndrome; type 1 and type 2 diabetes; end-stage kidney disease; motor neuron disease, multiple sclerosis, myasthenia gravis, or Huntington’s disease.

Our earlier review suggested a potentially large increase in severe outcomes among males versus females, and for people with some pre-existing conditions (heart failure, dementia, liver disease) [11]. The evidence from the present review no longer supports this increased risk, underscoring the importance of interpreting low certainty evidence cautiously, and of continually reviewing emerging data to support vaccine prioritization guidance. Our update found relatively strong evidence for little-to-no increase in severe outcomes for several pre-existing diseases, notably many cardiovascular and respiratory conditions. In contrast, many jurisdictions have identified people with these conditions as priorities for vaccination [16–19]. The reason for this is not fully clear, because the evidence on which prioritization decisions are made is often not publically available, and such decisions are informed by multiple considerations (e.g., local epidemiology, ethics, risk-benefit analyses, vaccine characteristics and availability [6, 7, 156]) and varying levels of evidence.

A large reliance on unadjusted data may provide some explanation [157]. The data synthesized within this review have the advantage of accounting for the impact of multiple co-existing conditions, social and demographic factors. For example, in a large study (n=418,794) of the general population in the United Kingdom [114], a large increase in hospitalization among people with heart failure (OR 2.24) and chronic obstructive pulmonary disease (OR 2.67) was observed. Following adjustment for age, sex, social factors, BMI, and other chronic conditions, these associations were substantially attenuated to the range of little-to-no association (aOR 1.09 and 1.51, respectively) [114]. We observed similar relationships in other studies. The thresholds of magnitude used to define ‘large’ and ‘very large’ increases in severe outcomes may also impact the conditions prioritized. Finally, there are some rare conditions (e.g., thalassemia, splenic dysfunction) for which to our knowledge, no large and well-adjusted studies have been published. In these cases, governing bodies may need to rely on evidence available for other infectious diseases, or from smaller cohorts suggesting the potential for increased risk [19, 158–161].

The findings of this review, among other considerations, have informed NACI’s vaccine prioritization guidance. Though access to COVID-19 vaccines is now widespread in Canada, this review continues to serve an important archival function. Our findings, particularly the developments that occurred between our first review and the present one, underscore the importance of continuing to review emerging data to provide up-to-date evidence of the highest possible certainty in times when urgent decision-making is required (e.g., pandemics, emergencies). As an exemplar, at the time of our first review, few large, well-adjusted analyses were available for children, who were not a population of high priority at that time. However, in our latest update in April 2021, we located six new eligible studies that helped NACI to develop evidence-informed guidance [32, 52, 55, 106, 123, 152]. Using the adapted methodology described herein, we were able to provide timely empirical evidence to inform decision-making without compromising aspects of rigour that were important to relevant stakeholders. This review may therefore serve as an exemplar for abbreviated methodologies that may be undertaken to inform decision-making in urgent situations.

## Limitations

Because we used a rapid approach, there is a possibility of undetected errors in study selection or data extraction. We mitigated this through piloting and engaging experienced reviewers. It was not appropriate to pool the findings due to substantial heterogeneity and at least some overlap in populations across studies; thus, it is possible that the qualitative estimates of measures of association across studies may have been over- or underestimated. Though the findings are most applicable to OECD countries, we often relied on data largely from the United States and other countries without universal healthcare systems. We located no evidence relevant to long-term outcomes that met our selection criteria.

## Conclusion

There is strong evidence to support at least a large increase in mortality from COVID-19 among older adults aged 60 to 69 years versus <60 years; people having two or more versus no comorbidities; and for people affected by Down syndrome; type 1 and 2 diabetes; end-stage kidney disease; and motor neuron disease, multiple sclerosis, myasthenia gravis, or Huntington’s disease (as a grouping). The methodology employed in this rapid systematic review may provide an important exemplar for future syntheses undertaken under urgent timelines.

## Data Availability

The data pertaining to the conclusions are available on reasonable request using the following link: https://doi.org/10.7939/DVN/W1M6OP.

## ADDITIONAL FILES

### Name: Additional file 1

Format: .docx

Title: Search strategy

Description: Search strategy used to locate records for the systematic review

### Name: Additional file 2

Format: .docx

Title: Approach to synthesis and all results

Description: Approach to data synthesis and full results for all populations analyzed

### Name: Additional file 3

Format: .docx

Title: Detailed study characteristics

Description: Characteristics of the included studies

## LIST OF ABBREVIATIONS

aOR: adjusted odds ratio
BMI: body mass index
COVID-19: novel coronavirus disease 2019
ICU: intensive care unit
RT-PCR: reverse transcription polymerase chain reaction
NACI: National Advisory Committee on Immunization
OECD: Organization for Economic Cooperation and Development

## DECLARATIONS

### Ethics approval and consent to participate

Not applicable.

### Consent for publication

Not applicable.

## Competing interests

The authors have no competing interests to declare. Michelle Gates and Allison Gates are employees of the Canadian Agency for Drugs and Technologies in Health (CADTH). The current work was unrelated to their employment and CADTH had no role in the funding, design, or oversight of the work.

## Funding

This study was completed under contract to the Public Health Agency of Canada, contract #4600001536. LH is supported by a Tier 1 Canada Research Chair in knowledge synthesis and translation. The funders had no role in the design of the study, data collection and analysis, preparation of the manuscript, nor decision to submit for publication.

## Authors’ contributions

MG contributed to the conception and design of the work, selection of studies, data extraction, analysis and interpretation of data, and drafted the initial version of the manuscript. JP contributed to the conception and design of the work, analysis and interpretation of data, and revised versions of the manuscript. AW, SG, SR, BZ, and AG contributed to the selection of studies, data extraction, organizing data for analysis, and revised versions of the manuscript. LH contributed to the conception and design of the work, methodological oversight, interpretation of findings, and revised versions of the manuscript. All authors approved of the final version of the manuscript submitted and agree to be accountable for all aspects of the work.

## Acknowledgements

We thank Kelsey Young, Kelly Farrah, Shainoor Ismail, and Matthew Tunis from the NACI Secretariat for expert input into the review’s scope and eligibility criteria, and for reviewing the initial draft of the manuscript; Sharon Bartholomew, Louise McRae, Anne-Marie Robert, Samina Aziz, Alain Demers, Catherine Pelletier, Cynthia Robitaille, Larry Shaver, Helene Gardiner, and Dianne Zakaria from the Public Health Agency of Canada for input into the necessary adjustment variables for analyses of chronic diseases; Elizabeth Dennett and Erica Wright for developing and running literature searches; Andrew Beck for assistance with screening; and Mackinna Hauff for article retrieval and document formatting.

## Notes

### Funding Statement

TThis study was completed under contract to the Public Health Agency of Canada, contract #4600001536. LH is supported by a Tier 1 Canada Research Chair in knowledge synthesis and translation. The funders had no role in the design of the study, data collection and analysis, preparation of the manuscript, nor decision to submit for publication.

### Author Declarations

Ethics approval was not required because this is a systematic review.

### Summary of Updates

Minor edits based on comments received from reviewers.

