## Additional file for "Risk factors associated with severe outcomes of COVID-19: A systematic rapid review to inform national guidance on vaccine prioritization in Canada"

**Appendix 1.** Search strategy

**A. Search strategy used to identify relevant studies on the magnitude of association between risk factors and severe outcomes of COVID-19**

| | **Stage 1 search for primary studies** | **Stage 2 search for systematic reviews** |
|---|---|---|
| **Approach** | Integration of studies from our team's previous review; updated search using strategies with the highest yield in the original review, and with modifications to include new (long-term) outcomes not included in the original review | Targeted searches when no primary studies on a risk factor of importance to NACI were located |
| **Strategy** | Shown in Additional file 1, part B | Shown in Additional file 1, parts C and D |
| **Concepts** | Cohort studies, COVID-19, $P^2$ROGRESS And Other Factors, severe short- and long-term outcomes | As in stage 1, with inclusion of terms for systematic reviews and specific conditions of interest: sickle cell anemia, thalassemia, cystic fibrosis, asplenia, learning disability |
| **Databases (date)** | Ovid Medline® ALL 1946- ; Epistemonikos COVID-19 in L-OVE Platform (https://app.iloveevidence.com/loves) for systematic reviews and broad syntheses categorized as reporting on individual predictors of outcomes (2-3 December 2020; updated in Medline for autoimmune conditions, immune compromise, and children 9-13 April 2021) | Ovid Medline® ALL 1946- (18 February 2021) |
| **Other sources (date)** | Websites: Government of Canada's First Nations and Inuit Health Branch, Public Health Agency of Canada, Public Health Ontario ICES, United States Centers for Disease Control and Prevention, Public Health England, Johns Hopkins Center for Health Security, European Centre for Disease Prevention and Control, GenderSci Lab COVID Project. [a] (6 January – 3 February 2021; updated for autoimmune conditions, immune compromise, and children 22 April-14 May 2021) | Not applicable |
| **Limits applied** | English and French language; 1 January 2020 to date of search; Organization for Economic Cooperation and Development countries | English and French language; 1 January 2020 to date of search |

NACI: National Advisory Committee on Immunization

[a] Selection of websites informed by NACI and the High Consequence Infectious Disease Working Group

**B. Main search for primary studies**

**Database:** Ovid MEDLINE(R) ALL 1946 to December 01, 2020
**Date searched:** December 2, 2020; updated 9-13 April 2021 for autoimmune conditions, immune compromise, and pediatrics

| | |
|---|---|
| 1 | (Risk factor* or relative risk or odds ratio or between group* or Regression or multi-variate or multivaria* or covariate or univariate or co-variate or matching or ANOVA or Analysis of variance or ANCOVA or Correlation or Covariance or Principal Component Analysis or cohort* or follow-up or prognos* or predict*).mp. |
| 2 | exp cohort studies/ or (cohort* or longitudinal or follow-up).mp. |
| 3 | (clinical data or clinical outcomes or (clinical adj5 (characteristics or features or manifestations))).tw,kf. |
| 4 | Transsexualism/ or bisexuality/ or exp homosexuality/ or exp disabled persons/ or transgendered persons/ or "transients and migrants"/ or exp refugees/ or exp Cultural Diversity/ or exp Socioeconomic Factors/ or (Metis or Indigenous* or Aboriginal* or Amerindian* or Autochton* or First Nations or First Nation or Inuit or Innu or Inuk or Inuvialuit or (native* adj3 (Canadian or American or Alaska)) or tribal or underprivileged or underrepresented or disadvantaged or disadvantage?ness or disparity or disparities or inequit* or inequalities or deprived or deprivation or minority or minorities or migrant* or immigrant* or visual* impair* or hearing impair* or amputat* or paraplegic* or quadraplegic* or wheelchair* or transsexual* or homosexual* or bisexual* or two spirit* or gender-questioning or HIV positive or "living with HIV" or shut-in or house-bound or neglected or (battered adj3 (spouse* or wife* or partner)) or disabled or "with a disability" or "with disabilities" or transgender* or poverty or impoverished or working poor or unemploy* or under-employed or low* soci* status or low* socioeconomic* or low* socio-economic* or low income* or low-SES or hard-to-house or homeless* or under-housed or (street adj4 (worker* or people or child or children or youth)) or (sex adj3 worker*) or prostitut* or inner city or downtown core or city core or skid row or alcoholic* or mental* ill* or mental disorder* or ((drug or substance*) adj3 (abuse* or use* or illegal or illicit or addict*))).mp. |
| 5 | 1 or 2 or 3 or 4 |
| 6 | (Mortal* or fatal* or death* or died or discharged alive or poor prognos* or good prognos* or clinical outcome* or adverse outcome* or disease course or clinical course or ((severe* or serious* or critical*) adj4 (ill* or outcome* or course or case or cases or patient* or condition)) or Severity or ((ICU or hospital or intensive care) adj7 (admission* or admit*)) or Ventilator* or ventilation or Hospitaliz* or hospitalis* or (Length adj3 stay)).mp. |
| 7 | (Quality of life or QOL or HRQoL or EDQ5 or EQ-5D or SF-36 or SF36 or SF-12 or SF12 or longhaul* or long-haul or ((longterm or long-term) adj4 (consequences or symptoms or sequelae or disabilit* or function*)) or stroke or ((lung* or kidney* or cardiac or heart) adj3 injur*) or functional status or post-discharge or ("after" adj3 discharge*) or ((function* or disability) adj5 (questionnaire* or self-report* or score or index or indices))).mp. |
| 8 | ((pregnan* or maternal or perinatal or birth or neonat* or infant*) adj7 outcome*).mp. |
| 9 | 6 or 7 or 8 |
| 10 | (Coronavirus* or corona-virus* or betacoronavirus* or nCOV* or 2019nCoV or 2019-ncov or covid or covid19 or SARS-CoV*or SARSCov*).mp. |
| 11 | (exp China/ or Iran/ or exp Russia/ or India/ or Pakistan/ or Bangladesh/ or Argentina/ or exp Africa/) not (canada/ or exp united states/ or europe/ or austria/ or belgium/ or exp france/ or exp germany/ or exp united kingdom/ or exp italy/ or spain/ or netherlands/ or exp "scandinavian and nordic countries"/ or australia/ or new zealand/ or mexico/ or chile/ or colombia/ or exp japan/ or korea/ or exp "republic of korea"/ or baltimore/ or berlin/ or boston/ or chicago/ or "district of columbia"/ or london/ or los angeles/ or new orleans/ or new york city/ or paris/ or philadelphia/ or rome/ or san francisco/ or estonia/ or latvia/ or lithuania/ or czech republic/ or hungary/ or poland/ or slovakia/ or slovenia/ or greece/ or luxembourg/ or portugal/ or switzerland/ or israel/ or turkey/) |
| 12 | ((China or wuhan or hubei or beijing).tw,kf. and china.in.) not (canada or italy or italian or spain or spanish or france or french or united kingdom or UK or england or english or NHS or ireland or irish or wales or welsh or scotland or scottish or german* or austria* or sweden or swedish or netherlands or norwegian or norway or finland or finnish or denmark or danish or european or belgium or belgian or Czech or Estonia* or Greece or |

| | |
|---|---|
| | Greek or Hungar* or Latvia* or Lithuania* or Luxembourg or Iceland* or Poland or Portugal or Slovak Republic or Slovenia* or Switzerland or Japan* or Tokyo or Korea* or Seoul or Chile* or Colombia* or Mexico or Mexican or Israel* or Turkey or Turkish or australia* or new zealand* or united states or USA or american or "U.S." or new york or california* or washington or seattle).tw,kf. |
| 13 | ((russia* or iran* or tehran or brazil* or India or Pakistan or Argentin* or South Africa* or Nigeria* or Morocc* or Ethiopia*) not (canada or italy or italian or spain or spanish or france or french or united kingdom or UK or england or english or NHS or ireland or irish or wales or welsh or scotland or scottish or german* or austria* or sweden or swedish or netherlands or norwegian or norway or finland or finnish or denmark or danish or european or belgium or belgian or Czech or Estonia* or Greece or Greek or Hungar* or Latvia* or Lithuania* or Luxembourg or Iceland* or Poland or Portugal or Slovak Republic or Slovenia* or Switzerland or Japan* or Tokyo or Korea* or Seoul or Chile* or Colombia* or Mexico or Mexican or Israel* or Turkey or Turkish or australia* or new zealand* or united states or USA or american or "U.S." or new york or california* or washington or seattle)).tw,kf. |
| 14 | ((china or russia or iran or tehran or Brazil or India or Pakistan or Argentin* or South Africa* or Nigeria* or Morocc* or Ethiopia*) not (canada or italy or italian or spain or spanish or france or french or united kingdom or UK or england or english or NHS or ireland or irish or wales or welsh or scotland or scottish or german* or austria* or sweden or swedish or netherlands or norwegian or norway or finland or finnish or denmark or danish or european or belgium or belgian or Czech or Estonia* or Greece or Greek or Hungar* or Latvia* or Lithuania* or Luxembourg or Iceland* or Poland or Portugal or Slovak Republic or Slovenia* or Switzerland or Japan* or Tokyo or Korea* or Seoul or Chile* or Colombia* or Mexico or Mexican or Israel* or Turkey or Turkish or australia* or new zealand* or united states or USA or american or "U.S." or new york or california* or washington or seattle)).in. |
| 15 | (intervention* or therap* or treatment* or management).ti. and (review or trial or guidelines).ti,pt. |
| 16 | (case reports/ or (case-stud* or case-report*).jw. or (case-study or (case-report not case-report form*)).mp.) not (case-series or case-control).mp. |
| 17 | (5 and 9 and 10) not (11 or 12 or 13 or 14 or 15 or 16) |
| 18 | limit 17 to yr="2020 -Current" |
| 19 | limit 18 to (english or french) |
| 20 | limit 19 to (comment or editorial or historical article or news or newspaper article) |
| 21 | 19 not 20 |

**Database:** Epistemonkos COVID-19 in L-OVE Platform, Inception-December 3, 2020
**Date searched:** December 3, 2020

Included studies categorized as “individual predictors of outcome”.

**C. Initial scoping search for systematic reviews**

**Database:** Ovid MEDLINE(R) ALL 1946 to Sept 11, 2020
**Date searched:** September 12, 2020

| | |
|---|---|
| 1 | (Risk factor* or relative risk or odds ratio or between group* or Regression or multi-variate or multivaria* or covariate or univariate or co-variate or matching or ANOVA or Analysis of variance or ANCOVA or Correlation or Covariance or Principal Component Analysis or cohort* or follow-up or prognos* or predict*).mp. |
| 2 | exp cohort studies/ or cohort*.mp. |
| 3 | ("Associated with" or "Association of" or "impact of" or "Correlated with" or "Impact* on" or characteristics or characterise or features or clinical findings or clinical outcomes or clinical manifestations or clinical course).ti. |
| 4 | (clinical data or (clinical adj5 (characteristics or features or manifestations))).tw,kf. |
| 5 | 1 or 2 or 3 or 4 |
| 6 | (Mortal* or fatal* or death* or died or discharged alive or poor prognos* or good prognos* or clinical outcome* or adverse outcome* or disease course or clinical course or ((severe* or serious* or critical*) adj4 (ill* or outcome* or course or case or cases or patient* or condition)) or Severity or ((ICU or hospital or intensive care) adj7 (admission* or admit*)) or Ventilator* or ventilation or Hospitaliz* or hospitalis* or (Length adj3 stay)).mp. |
| 7 | ((pregnan* or maternal or perinatal or birth or neonat* or infant*) adj7 outcome*).mp. |
| 8 | 6 or 7 |
| 9 | 5 and 8 |
| 10 | (Coronavirus* or corona-virus* or betacoronavirus* or nCOV* or 2019nCoV or 2019-ncov or covid or covid19 or SARS-CoV*or SARSCov*).mp. |
| 11 | limit 10 to yr="2020 -Current" |
| 12 | limit 11 to abstracts |
| 13 | (11 not 12) and (1 or 2 or 3 or 4 or 6 or 7) |
| 14 | 9 and 11 |
| 15 | 13 or 14 |
| 16 | (pubmed or medline or cochrane or scopus or cinahl).tw. or ((systematic* or evidence-based or scoping or umbrella or rapid) adj3 (review* or overview*)).pt,mp,jw. or meta-analy*.pt,mp. or (meta-analy* or metaanalys* or research-synthesis).tw. or search*.ab. or (hta or technology assessment).mp,jw. |
| 27 | 15 and 16 |
| 18 | limit 17 to (english or french) |

**Database:** Epistemonkos COVID-19 in L-OVE Platform, Inception-December 3, 2020
**Date searched:** September 12, 2020

Included reviews and broad syntheses categorized as “individual predictors of outcome”.

### D. Targeted searches for evidence gaps

**Database:** Ovid MEDLINE(R) ALL 1946 to February 17, 2021
**Data searched:** February 18, 2021

| | |
|---|---|
| 1 | (Coronavirus* or corona-virus* or betacoronavirus* or nCOV* or 2019nCoV or 2019-ncov or covid or covid19 or SARS-CoV*or SARSCov*).mp. (118913) |
| 2 | (Risk factor* or relative risk or odds ratio or between group* or Regression or multi-variate or multivaria* or covariate or univariate or co-variate or matching or ANOVA or Analysis of variance or ANCOVA or Correlation or Covariance or Principal Component Analysis or cohort* or follow-up or prognos* or predict*).mp. (6114068) |
| 3 | exp cohort studies/ or cohort*.mp. (2391946) |
| 4 | ("Associated with" or "Association of" or "impact of" or "Correlated with" or "Impact* on" or characteristics or characterise or features or clinical findings or clinical outcomes or clinical manifestations or clinical course).ti. (1197372) |
| 5 | (clinical data or (clinical adj5 (characteristics or features or manifestations))).tw,kf. (405780) |
| 6 | (Mortal* or fatal* or death* or died or discharged alive or poor prognos* or good prognos* or clinical outcome* or adverse outcome* or disease course or clinical course or ((severe* or serious* or critical*) adj4 (ill* or disease* or outcome* or course or case or cases or patient* or condition)) or Severity or ((ICU or hospital or intensive care) adj7 (admission* or admit*)) or Ventilator* or ventilation or Hospitaliz* or hospitalis* or (Length adj3 stay)).mp. (3625824) |
| 7 | or/2-6 (9191992) |
| 8 | 1 and 7 (53864) |
| 9 | limit 8 to yr="2019 -Current" (49600) |
| 10 | limit 9 to (english or french) (48382) |
| 11 | exp Anemia, Sickle Cell/ (22908) |
| 12 | sickle cell.ti,ab,kf. (25749) |
| 13 | or/11-12 (29702) |
| 15 | 10 and 13 (45) **42 without duplicates** |
| 16 | exp Thalassemia/ (23108) |
| 16 | thalassemi*.ti,ab,kf. (18243) |
| 17 | or/15-16 (28112) |
| 18 | 10 and 17 (24) **18 without duplicates** |
| 19 | Cystic Fibrosis/ (35986) |
| 20 | (cystic fibrosis or fibrocystic).ti,ab,kf. (48131) |
| 21 | or/19-20 (53420) |
| 22 | 10 and 21 (58) **55 without duplicates** |
| 23 | Splenectomy/ (21856) |
| 24 | (aspleni* or hyposplenI* or splenectom*).ti,ab,kf. (25463) |
| 25 | or/23-24 (33334) |
| 26 | 10 and 25 (10) **7 without duplicates** |
| 27 | learning disabilities/ or exp intellectual disability/ (110479) |
| 28 | ((intellectual* or learning or developmental) adj2 disab*).ti,ab,kf. (33796) |
| 29 | 27 or 28 (126771) |
| 30 | 10 and 29 (54) no duplicates |
| 31 | 14 or 18 or 22 or 26 or 30 (180) |

**Appendix 2.** Details of the synthesis approach and summary of all findings from the updated rapid review

**A. Approach to synthesis and drawing conclusions**

| Stage of review | Approach and considerations |
|---|---|
| Synthesis | 1. Estimated the magnitude of association based on the studies contributing the most events (or largest sample size if events unavailable)<br><br>2. Compared the findings to other studies based on relevancy (i.e., most well and suitably adjusted, taking place in universal healthcare countries)<br><br>3. Mapped and considered potential overlap across studies conducted in the same country<br><br>4. Two research leads reached consensus on the best estimate of association, and categorized the magnitude of association (informed by aOR, aRR, or aHR) as:<br>– Little-to-no: <2.0 for increase, >0.50 for reduction<br>– Large: 2.0 to 3.9 for increase, 0.50 to 0.26 for reduction<br>– Very large: ≥4.0 for increase, <0.25 for reduction |
| Drawing conclusions | Two research leads reached consensus on the certainty of each estimated association. All exposure-outcome comparisons started at high certainty. We rated down for:<br><br>1. **Risk of bias:** (a) lack of adjustment for social factors; (b) the expectation of attenuated magnitude of association when the model was adjusted for laboratory values and/or symptoms; (c) for the hospitalization outcome, potential for testing bias in certain populations (e.g., healthcare providers), where the exposure group may have had less severe disease than those not exposed; (d) use of historical, potentially inaccurate, data for risk factors or covariates.<br><br>2. **Inconsistency:** for example, different magnitude of association across studies or populations; inadequate number of studies to demonstrate consistency.<br><br>3. **Imprecision:** for example, small sample size; one study with a wide confidence interval indicating the possibility of two or more different conclusions.<br><br>4. **Indirectness:** for example, all studies from countries without universal healthcare; inclusion of conditions that may not be directly applicable (e.g., need to extrapolate from different age groups than that of primary interest). |

aHR=adjusted hazard ratio; aOR=adjusted odds ratio; aRR=adjusted risk ratio

**B. Summary of findings for multivariate-adjust associations**

**Table A2.1.** Summary of findings for multivariate-adjusted associations between risk factors and severe outcomes of COVID-19 among the general population

| Risk factor vs. comparator | **Magnitude of association (certainty in association), by outcome** Magnitude of associations are shown as: uncertain (very low certainty), little-to-no association (-; ≤2.0), large association (+; 2.0 to 3.0), very large association (++; >3.0). | |
|---|---|---|
| | **Hospitalization** | **Mortality** |
| **Age & sex/gender** | | |
| 60-69 years vs. <60 years | + (low) | + (moderate) / ++ (low) |
| Male vs. female | - (low) | - (moderate) |
| **High risk of exposure due to occupation** | | |
| Healthcare worker vs. non-healthcare worker | - (low) | |
| Patient facing healthcare worker vs. non-patient facing healthcare worker | + (low) | |
| Household member of patient-facing vs. non-patient facing healthcare worker | uncertain | |
| **Immunocompromised: cancer or cancer treatment (present vs. absent)** | | |
| Malignancy | | - (moderate) |
| Chemotherapy in the past 12 months (Grades A-C) [a] | | + / ++ (low) |
| Radiotherapy in the past 6 months | | + (low) |
| **Immunocompromised: transplant (present vs. absent)** | | |
| Recent bone marrow or stem cell transplant | | + (low) |
| Solid organ transplant, excluding kidney | | - (low) |
| Kidney transplant (due to stage 5 kidney disease) | | + (low) |
| **Immunocompromised: immunosuppression or immunodeficiency (present vs. absent)** | | |
| Immunodeficiency or immunosuppression (unspecified) | - (moderate) | - (moderate) |
| Sickle cell disease or severe immunodeficiency | | + (low) |
| **Autoimmune conditions (present vs. absent)** | | |
| Rheumatoid arthritis or systemic lupus erythematosus | | - (moderate) |
| Inflammatory rheumatic diseases (any) | - (moderate) | |
| Rheumatoid arthritis | - (moderate) | |
| Spondyloarthritis | - (low) | |
| Connective tissue disease | - (low) | |
| Vasculitis | + (low) | |
| **Pre-existing conditions: number of comorbidities** | | |
| 1 vs. no comorbidities | + (low) | |
| 2 or more vs.no comorbidities | ++ (low) | |
| **Pre-existing conditions: underweight, overweight, and obesity** | | |
| Underweight vs. normal weight | - (low) | |
| Overweight vs. normal weight | - (low) | |
| Obesity (all classes) vs. normal weight | + (low) | |
| Obesity class I vs. normal weight | - (low) | |
| Obesity class II or III vs. normal weight | + (low) | |
| **Pre-existing conditions: cardiovascular (present vs. absent)** | | |

| Risk factor vs. comparator | **Magnitude of association (certainty in association), by outcome** Magnitude of associations are shown as: uncertain (very low certainty), little-to-no association (-; ≤2.0), large association (+; 2.0 to 3.0), very large association (++; >3.0). | |
|---|---|---|
| | **Hospitalization** | **Mortality** |
| Congenital heart disease | | - (moderate) |
| Cardiovascular disease | - (low) | |
| Coronary artery disease | - (low) | - (moderate) |
| Peripheral vascular disease | | - (moderate) |
| Hypertension | - (low) | |
| Atrial fibrillation | | - (moderate) |
| Stroke | - (low) | - (moderate) |
| Thromboembolism | | - (moderate) |
| Congestive heart failure | | - (moderate) |
| Heart failure | - (low) | |
| **Pre-existing conditions: respiratory (present vs. absent)** | | |
| History of pneumonia | - (low) | |
| Asthma | | - (moderate) |
| Chronic obstructive pulmonary disease | - (low) | - (moderate) |
| Pulmonary hypertension or fibrosis | | - (low) |
| Bronchiectasis, cystic fibrosis, or alveolitis | | - (moderate) |
| **Pre-existing conditions: endocrine (present vs. absent)** | | |
| Type 1 diabetes | | + (moderate) / ++ (low) |
| Type 2 diabetes | | + (moderate) / ++ (low) |
| Diabetes (any) | - (low) | |
| **Pre-existing conditions: renal (present vs. absent)** | | |
| Chronic kidney disease | + (low) | - (moderate) |
| End-stage kidney disease | | + (moderate) |
| End-stage kidney disease with dialysis | | + (moderate) / ++ (low) |
| **Pre-existing conditions: hepatic (present vs. absent)** | | |
| Cirrhosis | | - (low) |
| **Pre-existing conditions: neurological (present vs. absent)** | | |
| Alzheimer's disease or dementia | uncertain | |
| Dementia | | + (moderate) |
| Parkinson's disease | | + (low) |
| Epilepsy | | - (moderate) |
| Motor neuron disease, multiple sclerosis, myasthenia gravis, or Huntington's disease | | + (moderate) |
| Cerebral palsy | | + (low) |
| **Pre-existing conditions: other (present vs. absent)** | | |
| Down Syndrome | + (moderate) / ++ (low) | ++ (moderate) |
| Intellectual disability (excluding Down Syndrome) | | - (moderate) |
| Osteoporotic fracture (hip, spine, wrist) | | - (moderate) |
| Severe mental illness | | - (moderate) |
| **Frailty (present vs. absent)** [b] | | |
| Pre-frailty | - (low) | |
| Frailty | + (low) | |
| **Other factors** | | |
| Past or current vs. never smoking | - (low) | |
| Higher vs. lower alcohol consumption | - (low) | |

| Risk factor vs. comparator | **Magnitude of association (certainty in association), by outcome** Magnitude of associations are shown as: uncertain (very low certainty), little-to-no association (-; ≤2.0), large association (+; 2.0 to 3.0), very large association (++; >3.0). | |
|---|---|---|
| | **Hospitalization** | **Mortality** |
| Low vs. adequate physical activity | - (low) | |

[a] There was evidence of a large to very large increase in mortality with grades B and C chemotherapy from one large study. However, the stages of chemotherapy were not defined, and could not be ascertained from the study's authors. In the absence of adequate information, we have grouped all stages of chemotherapy (A, B, and C) together for analysis.

[b] General population sample that may include community and non-community dwelling people. Measured on scales that include items such as weight loss, exhaustion, physical activity, walking speed, grip strength, overall health, disability, presence of disease, dementia, falls, mental wellbeing.

**Table A2.2.** Summary of findings for multivariate-adjusted associations between risk factors and severe outcomes of COVID-19 among people with laboratory-confirmed COVID-19

| Risk factor vs. comparator | **Magnitude of association (certainty in association), by outcome**<br>Magnitude of associations are shown as: uncertain (very low certainty), little-to-no association (-; ≤2.0), large association (+; 2.0 to 3.0), very large association (++; >3.0). | | | | |
|---|---|---|---|---|---|
| | **Hospitalization** | **ICU admission** | **Mechanical ventilation** | **Severe disease** | **Mortality** |
| **Age & sex/gender** | | | | | |
| 60-69 years vs. <60 years | + (moderate) | uncertain | + (low) | + to ++ (low) | + (moderate) / ++ (low) |
| Children (0-18 years) vs. adults (18-44 years) | - (low) | | | - (low) | |
| Children (0-19 years) vs. adults (50-59 years) | + reduction (low) | | | | |
| Children <1 month vs. >1 month | | - (low) | | | |
| **Sex or gender identity** | | | | | |
| Male vs. female | - (moderate) | - (low) | - (low) | - (moderate) | - (high) |
| Among children: male vs. female | - (low) | - (low) | - (low) | - (low) | |
| **Among children, race or ethnicity** | | | | | |
| Black vs. White | | | - (low) | | |
| Black vs. White non-Hispanic | + (low) | | | + (low) | |
| Hispanic/Latino vs. White non-Hispanic | - (low) | | | - (low) | |
| Asian vs. White non-Hispanic | - (low) | | | - (low) | |
| Hawaiian/Pacific Islander vs. White non-Hispanic | - (low) | | | - (low) | |
| Alaskan/American Indian vs. White non-Hispanic | - (low) | | | - (low) | |
| **Pregnancy (present vs. absent)** | | | | | |
| Pregnancy (any stage) | + (low) | + (low) | + (low) | | - (low) |
| **High risk of exposure due to occupation** | | | | | |
| Healthcare worker vs. non-healthcare worker | + reduction (low) | + reduction (low) | | | |
| Patient facing healthcare worker vs. non-patient facing healthcare worker | uncertain | uncertain | | | |
| **Immunocompromised (present vs. absent)** | | | | | |
| Malignancy (any type, severity, or stage) | - (moderate) | - (low) | - (low) | - (moderate) | - (low) |
| Metastatic cancer among those age <70 years | | | | | + (low) |
| Active cancer treatment | uncertain | uncertain | uncertain | | uncertain |
| History of cancer without current treatment | - (low) | - (low) | - (low) | | - (low) |

| Risk factor vs. comparator | Magnitude of association (certainty in association), by outcome<br>Magnitude of associations are shown as: uncertain (very low certainty), little-to-no association (-; ≤2.0), large association (+; 2.0 to 3.0), very large association (++; >3.0). | | | | |
|---|---|---|---|---|---|
| | Hospitalization | ICU admission | Mechanical ventilation | Severe disease | Mortality |
| Solid organ transplant | | | | | + (low) |
| Transplant (any type) | + (low) | | | | |
| Human immunodeficiency virus | uncertain | | | uncertain | - (moderate) |
| Immunosuppression (unspecified) | - (moderate) | | | uncertain | - (low) |
| Among children, immunodeficiency or immunosuppression | + (low) | - (low) | uncertain | + to ++ (low) | |
| Among children, malignancy | | | + (low) | | |
| **Autoimmune conditions (present vs. absent)** | | | | | |
| Autoimmune diseases (general) | - (low) | | | | |
| Rheumatic diseases and connective tissue disorders [a] | - (low) | - (low) | - (low) | - (moderate) | - (moderate) |
| Inflammatory bowel disease | - (low) | - (low) | | - (low) | - (low) |
| Multiple sclerosis | | | | uncertain | |
| **Pre-existing conditions: number of comorbidities** | | | | | |
| 1 vs. no comorbidities | - (moderate) | uncertain | uncertain | | - (low) |
| 2 or more vs. no comorbidities | + (moderate) | + (low) | + (low) | | + (moderate) |
| Among children, 1 vs. no comorbidities [b] | + (low) | | | | |
| Among children, 1 or more vs. no comorbidities [c] | | + (low) | + (low) | + (low) | |
| Among children, 2 or more vs. no comorbidities [b] | + (moderate) | | | | |
| **Pre-existing conditions: underweight, overweight, and obesity** | | | | | |
| Underweight vs. normal weight | uncertain | | uncertain | | - (low) |
| Underweight vs. normal weight, overweight, or class I obesity | | | | | - (low) |
| Overweight vs. normal weight | - (moderate) | | - (low) | - (low) | - (moderate) |
| Obesity class I vs. normal weight | | | | | - (moderate) |
| Obesity class II vs. normal weight | | | | | - (low) |
| Obesity class I or II vs. normal weight | - (moderate) | | | | |
| Obesity class III vs. normal weight | - (low) | | | | + (low) |
| Obesity (all classes) vs. normal weight | | | - (low) | - (low) | |
| Obesity (all classes); present vs. absent | - (moderate) | | | - (low) | - (high) |
| Obesity class III; present vs. absent | + (low) | | | | |
| Among children, obesity | uncertain | - (low) | | uncertain | |
| **Pre-existing conditions: cardiovascular (present vs. absent)** | | | | | |

| Risk factor vs. comparator | **Magnitude of association (certainty in association), by outcome** Magnitude of associations are shown as: uncertain (very low certainty), little-to-no association (-; ≤2.0), large association (+; 2.0 to 3.0), very large association (++; >3.0). | | | | |
|---|---|---|---|---|---|
| | **Hospitalization** | **ICU admission** | **Mechanical ventilation** | **Severe disease** | **Mortality** |
| Atrial fibrillation | - (low) | | | | |
| Cardiovascular disease | - (low) | | | | - (low) |
| Cerebrovascular accident | + (moderate) | | | | |
| Cerebrovascular disease | - (moderate) | | | - (low) | - (moderate) |
| Congestive heart failure or heart failure | - (low) | | | | - (moderate) |
| Coronary artery disease | - (high) | | | | - (moderate) |
| Hyperlipidemia | - (moderate) | | | | - (moderate) |
| Hypertension | - (high) | | | - (low) | - (high) |
| Myocardial infarction | | | | | - (low) |
| Peripheral vascular disease | - (low) | | | | - (low) |
| Venous thromboembolism | - (low) | | | | |
| Among children, cardiovascular disease | ? (uncertain) | | | - (low) | |
| Among children, hypertension | + (low) | | | - (low) | |
| **Pre-existing conditions: respiratory (present vs. absent)** | | | | | |
| Asthma | - (high) | | - (low) | | - (high) |
| Chronic obstructive pulmonary disease | - (high) | | - (low) | | - (high) |
| Chronic bronchitis | - (low) | | | | |
| Interstitial lung disease | uncertain | | | | |
| Obesity hypoventilation | - (low) | | + (low) | | - (low) |
| Rhinitis or rhinosinusitis | - (low) | | | | |
| Among children, asthma | - (low) | + (low) | | | |
| **Pre-existing conditions: endocrine (present vs. absent)** | | | | | |
| Diabetes (any) | - (low) [d] | | - (low) | - (low) | - (moderate) |
| Glycosylated hemoglobin <7.5 vs. ≥7.5% | | | | | - (low) |
| Diabetes (any) in females | | | | | + (low) |
| Hypothyroidism | - (low) | | | | |
| Among children, diabetes | ? (uncertain) | - (low) | | - (low) | |
| Among children, endocrine condition | | | + (low) | | |
| **Pre-existing conditions: renal (present vs. absent)** | | | | | |
| Kidney disease (any) | | | | | - (moderate) |
| Chronic kidney disease | - (low) [d] | | - (low) | | - (low) |

| Risk factor vs. comparator | **Magnitude of association (certainty in association), by outcome** Magnitude of associations are shown as: uncertain (very low certainty), little-to-no association (-; ≤2.0), large association (+; 2.0 to 3.0), very large association (++; >3.0). | | | | |
|---|---|---|---|---|---|
| | **Hospitalization** | **ICU admission** | **Mechanical ventilation** | **Severe disease** | **Mortality** |
| End-stage kidney disease | uncertain | | | | |
| Dialysis | | | - (low) | | - (low) |
| **Pre-existing conditions: hepatic (present vs. absent)** | | | | | |
| Cirrhosis | - (low) | | - (low) | | - (low) |
| **Pre-existing conditions: neurological (present vs. absent)** | | | | | |
| Alzheimer's disease or dementia | | | | - (low) | |
| Dementia | uncertain | | | | - (moderate) |
| Epilepsy | | | | - (low) | |
| Hemi- or paraplegia | | | | | - (moderate) |
| Parkinson's disease or movement disorder | | | | - (low) | |
| **Pre-existing conditions: psychiatric (present vs. absent)** | | | | | |
| Any mental illness/mood disorder | | | | - (moderate) | - (moderate) |
| Depression | - (low) | | | | |
| Major psychiatric disorder [e] | + (low) | | | - (low) | + (low) |
| **Pre-existing conditions: other (present vs. absent)** | | | | | |
| Disability | | | | | - (low) |
| Peptic ulcer | | | | | - (low) |
| Intellectual disability or developmental disorder | | | | - (low) | |
| Obstructive sleep apnea | - (moderate) | | - (low) | | - (low) |
| Among children, metabolic condition | | | + (low) | | |
| **Other factors** | | | | | |
| Past or current vs. never smoking | uncertain | | - (low) | | - (moderate) |
| Higher vs. lower alcohol consumption | - (low) | | | | |
| Alcohol abuse vs. no alcohol abuse | - (low) | | - (low) | | - (low) |
| Substance abuse vs. no substance abuse | uncertain | | | uncertain | uncertain |

ICU=intensive care unit

[a] There was also low certainty evidence of no increase in renal failure or ischemic stroke with rheumatic disease. These outcomes were not investigated for any other risk factors in this population.

[a] Includes congenital malformations, asthma, epilepsy, complex genetic syndromes, endocrine disorders, cancers, hematologic diseases, rheumatic diseases, autoimmune disease, autism or neurologic development impairment, gastrointestinal diseases, liver disease, renal disease, genitourinary diseases, cystic fibrosis or other chronic lung diseases, metabolic disorders, hydrocephalus, severe obesity, hypertension, otolaryngolic diseases, pregnancy, and 'complex chronic conditions'.

[b] Includes congenital malformations, asthma, epilepsy, complex genetic syndromes, endocrine disorders, cancers, hematologic diseases, rheumatic diseases, autism or neurologic development impairment, gastrointestinal diseases, genitourinary diseases, renal disease, cystic fibrosis or other chronic lung diseases, metabolic disorders, hydrocephalus, severe obesity, otolaryngolic diseases.

[c] Approaching a large association (OR 1.8-1.9)

[d] Schizophrenia, schizoaffective disorder, or bipolar disorder. Defined by hospital discharge diagnosis, in combination with drug use (filled a prescription) for the condition in the past 6 months.

**Table A2.3.** Summary of findings for multivariate-adjusted associations between risk factors and severe outcomes of COVID-19 among people hospitalized with laboratory-confirmed COVID-19

| **Risk factor vs. comparator** | **Magnitude of association (certainty in association), by outcome** Magnitude of associations are shown as: uncertain (very low certainty), little-to-no association (-; ≤2.0), large association (+; 2.0 to 3.0), very large association (++; >3.0). | | | | | |
|---|---|---|---|---|---|---|
| | **Hospital length of stay** | **ICU admission** | **Mechanical ventilation** | **Severe disease** | **Acute kidney injury** | **Mortality** |
| **Age & sex/gender** | | | | | | |
| 60-69 years vs. <60 years | uncertain | - (low) | - (low) | + (low) | - (moderate) | + (high) |
| Male vs. female | - (low) | - (moderate) | - (moderate) | - (moderate) | - (low) | - (high) |
| Among children, male vs. female | | | | - (low) | | |
| **Among children, race or ethnicity** | | | | | | |
| Hispanic or Latino vs. White non-Hispanic | | | | - (low) | | |
| Asian vs. White non-Hispanic | | | | uncertain | | |
| Black vs. White non-Hispanic | | | | - (low) | | |
| **High risk of exposure due to occupation** | | | | | | |
| Healthcare worker vs. non-healthcare worker | | + reduction (low) | | | | |
| **Immunocompromised (present vs. absent)** | | | | | | |
| Malignancy (any type, severity, or stage) [a] | | - (moderate) [a] | - (moderate) | - (moderate) | - (moderate) | - (moderate) [a] |
| Transplant (any type) | | - (low) | | | | |
| Solid organ transplant | | | - (low) | - (low) | + (low) | - (low) |
| Human immunodeficiency virus | | - (low) | | | | - (low) |
| Immunosuppression or immunodeficiency (unspecified) | | - (low) | | uncertain | uncertain | - (moderate) |
| **Autoimmune conditions (present vs. absent)** | | | | | | |
| Autoimmune disease (any type) | | uncertain | | | | |
| Connective tissue disorder | | | | | uncertain | |
| Rheumatic disease | | | | | | - (low) |
| Chronic inflammatory disease | | | - (low) | | | - (low) |
| Inflammatory bowel disease [b] | | | - (low) | | | - (low) |
| **Pre-existing conditions: number of comorbidities** | | | | | | |
| 1 vs. no comorbidities | - (low) | - (moderate) | - (low) | - (low) | | - (moderate) |
| 2 or more vs. no comorbidities | - (low) | - (low) | - (low) | + (low) | | + (moderate) |
| Among children, chronic condition (present vs. absent) | | | | + (low) | | ++ (low) |
| **Pre-existing conditions: underweight, overweight, and obesity** | | | | | | |
| Underweight vs. normal weight | | | uncertain | | | - (moderate) |

| Risk factor vs. comparator | Magnitude of association (certainty in association), by outcome<br>Magnitude of associations are shown as: uncertain (very low certainty), little-to-no association (-; ≤2.0), large association (+; 2.0 to 3.0), very large association (++; >3.0). | | | | | |
|---|---|---|---|---|---|---|
| | **Hospital length of stay** | **ICU admission** | **Mechanical ventilation** | **Severe disease** | **Acute kidney injury** | **Mortality** |
| Underweight vs. normal weight or overweight | | | | | | - (low) |
| Overweight vs. normal weight | | | - (low) | - (moderate) | | - (moderate) |
| Obesity class I vs. normal weight | | | uncertain | | | - (low) |
| Obesity class II vs. normal weight | | | uncertain | | | uncertain |
| Obesity class I or II vs. normal weight | | | | - (low) | | - (low) |
| Obesity class II or III vs. normal weight | | | | | | - (low) |
| Obesity class III vs. normal weight | | | uncertain | uncertain | | - (low) |
| Obesity (all classes) vs. normal weight | | | - (low) | | - (low) | - (low) |
| Obesity (all classes); present vs. absent | | - (low) | | | - (low) | - (high) |
| Obesity class I or II; present vs. absent | | | - (low) | | | |
| Obesity class III; present vs. absent | | | uncertain | + (low) | | |
| **Pre-existing conditions: cardiovascular (present vs. absent)** | | | | | | |
| Atrial fibrillation | | - (low) | - (low) | - (low) | | - (moderate) |
| Cardiomyopathy | | | | | | |
| Cardiovascular disease | | - (low) | | - (moderate) | - (moderate) | - (high) |
| Cerebrovascular accident | | | - (low) | - (low) | | - (moderate) |
| Cerebrovascular disease | | - (low) | | | - (low) | - (low) |
| Congestive heart failure or heart failure | | - (moderate) | - (low) | - (low) | | - (high) |
| Coronary artery disease | | - (moderate) | - (moderate) | - (low) | | - (high) |
| Hyperlipidemia | | - (low) | - (low) | - (low) | | - (moderate) |
| Hypertension | | - (moderate) | - (moderate) | - (moderate) | - (moderate) | - (high) |
| Myocardial infarction | | | | | | |
| Peripheral vascular disease | | | - (low) | - (low) | - (low) | - (moderate) |
| Venous thromboembolism | | | - (low) | - (low) | | - (low) |
| **Pre-existing conditions: respiratory (present vs. absent)** | | | | | | |
| Asthma | | - (low) | - (moderate) | - (moderate) | | - (high) |
| Asthma or chronic obstructive pulmonary disease | | | | | - (low) | |
| Chronic lung disease | | | | | | |
| Chronic obstructive pulmonary disease | | | - (moderate) | - (low) | | - (high) |
| Interstitial lung disease | | uncertain | | | | |
| Obstructive lung disease (any) | | - (moderate) | | | | |
| **Pre-existing conditions: endocrine (present vs. absent)** | | | | | | |
| Diabetes (any) | | - (moderate) | - (moderate) | - (moderate) | - (moderate) | - (high) |

| Risk factor vs. comparator | Magnitude of association (certainty in association), by outcome Magnitude of associations are shown as: uncertain (very low certainty), little-to-no association (-; ≤2.0), large association (+; 2.0 to 3.0), very large association (++; >3.0). | | | | | |
|---|---|---|---|---|---|---|
| | **Hospital length of stay** | **ICU admission** | **Mechanical ventilation** | **Severe disease** | **Acute kidney injury** | **Mortality** |
| Hypothyroidism | | | - (low) | - (low) | | - (low) |
| **Pre-existing conditions: renal (present vs. absent)** | | | | | | |
| Chronic kidney disease | | - (moderate) | - (moderate) | - (low) | | - (high) |
| End-stage kidney disease | - (low) | | - (low) | | | - (moderate) |
| **Pre-existing conditions: hepatic (present vs. absent)** | | | | | | |
| Liver disease or cirrhosis | | | | | | - (moderate) |
| Liver disease | | | | | - (low) | |
| **Pre-existing conditions: neurological (present vs. absent)** | | | | | | |
| Chronic neurologic disorder | | | | | | - (moderate) |
| Dementia | | | - (low) | uncertain | - (moderate) | - (low) |
| Neurologic disease | | - (low) | | | | |
| Paraplegia | | | | | - (low) | uncertain |
| **Pre-existing conditions: psychiatric (present vs. absent)** | | | | | | |
| Schizophrenia | | - (low) | | | | - (low) |
| **Pre-existing conditions: other (present vs. absent)** | | | | | | |
| Hematologic disorder | | | | | | - (low) |
| Obstructive sleep apnea | | - (low) | | | | - (low) |
| **Frailty** | | | | | | |
| Pre-frailty | - (low) | - (low) | - (low) | | | - (low) |
| Frailty | + (low); - (low) for stay >10 day | + (low) | - (low) | | | + (low) |
| **Other factors** | | | | | | |
| Past or current vs. never smoking | - (low) | - (low) | - (low) | - (moderate) | - (low) | - (moderate) |

ICU=intensive care unit

[a] Includes hematologic and lymphatic malignancy

[b] Includes inflammatory bowel disease, rheumatoid arthritis, spondyloarthritis, and psoriatic arthritis

**Table A2.4.** Summary of findings for multivariate-adjusted associations between risk factors and severe outcomes of COVID-19 among people admitted to the intensive care unit (all outcomes), or among people mechanically ventilated (mortality only) with laboratory-confirmed COVID-19

| Risk factor vs. comparator | **Magnitude of association (certainty in association), by outcome** Magnitude of associations are shown as: uncertain (very low certainty), little-to-no association (-; ≤2.0), large association (+; 2.0 to 3.0), very large association (++; >3.0). | | | | | |
|---|---|---|---|---|---|---|
| | **ICU length of stay** | **Mechanical ventilation** | **Severe disease** | **Acute kidney injury** | **Mortality (among ICU admitted)** | **Mortality (among mechanically ventilated)** |
| **Age & sex/gender** | | | | | | |
| 60-69 years vs. <60 years | | | - (low) | - (low) | - (low) | - (low) |
| Male vs. female | | uncertain | | uncertain | - (moderate) | - (low) |
| **Immunocompromised (present vs. absent)** | | | | | | |
| Active malignancy | | | | | | + (low) |
| Malignancy (any type, severity, or stage) | | uncertain | | - (low) | - (low) | - (low) |
| Transplant (solid organ) | | - (low) | | - (low) | - (low) | |
| Immunosuppression (unspecified) | | | | | - (low) | |
| **Autoimmune conditions (present vs. absent)** | | | | | | |
| Systemic inflammatory disease | | | | | - (low) | |
| **Pre-existing conditions: number of comorbidities** | | | | | | |
| 1 vs. no comorbidities | - (low) | | | | | |
| 2 or more vs.no comorbidities | - (low) | | | | | |
| **Pre-existing conditions: underweight, overweight, and obesity** | | | | | | |
| Underweight vs. normal weight | | | | | | uncertain |
| Overweight vs. normal weight | | | | uncertain | uncertain | - (low) |
| Obesity class I vs. normal weight | | | | uncertain | - (low) | uncertain |
| Obesity class II vs. normal weight | | | | uncertain | uncertain | - (low) |
| Obesity class III vs. normal weight | | | | + (low) | uncertain | - (low) |
| Obesity (all classes); present vs. absent | | | | | - (low) | |
| **Pre-existing conditions: cardiovascular (present vs. absent)** | | | | | | |
| Chronic heart disease | | | | | - (low) | |
| Coronary artery disease | | | | - (low) | - (low) | - (low) |
| Hypertension | | | | - (low) | - (moderate) | - (low) |
| Congestive heart failure (or heart failure for mortality among mechanically ventilated) | | | | - (low) | - (low) | - (low) |
| **Pre-existing conditions: respiratory (present vs. absent)** | | | | | | |

| Risk factor vs. comparator | Magnitude of association (certainty in association), by outcome<br>Magnitude of associations are shown as: uncertain (very low certainty), little-to-no association (-; ≤2.0), large association (+; 2.0 to 3.0), very large association (++; >3.0). | | | | | |
|---|---|---|---|---|---|---|
| | **ICU length of stay** | **Mechanical ventilation** | **Severe disease** | **Acute kidney injury** | **Mortality (among ICU admitted)** | **Mortality (among mechanically ventilated)** |
| Asthma | | | | | - (low) | - (low) |
| Chronic obstructive pulmonary disease | | | | | - (low) | - (low) |
| Chronic respiratory disease | | | | | - (low) | |
| **Pre-existing conditions: endocrine (present vs. absent)** | | | | | | |
| Diabetes (any) | | | | - (low) | - (moderate) | - (low) |
| **Pre-existing conditions: renal (present vs. absent)** | | | | | | |
| Chronic kidney disease, any stage | | | | | - (low) | - (low) |
| Chronic kidney disease, stage III | | | | - (low) | | |
| Chronic kidney disease, stage IV and V | | | | + (low) | | |
| End-stage kidney disease | | | | | | - (low) |
| **Pre-existing conditions: neurological (present vs. absent)** | | | | | | |
| Chronic neurologic disorder | | | | | - (low) | |
| **Other factors** | | | | | | |
| Past or current vs. never smoking | | | | | - (low) | - (low) |

ICU=intensive care unit

**C. Findings of Canadian reports**

**Table A2.5.** All findings from Canadian reports

| Outcome (population) | Study | Key findings |
|---|---|---|
| **Age 60-69 vs. <60 years** | | |
| Hospitalization (people with COVID-19) | O'Brien 2020 (a) | – The highest hospitalization rates were in those aged ≥70 years (about 40% in females and 50% in males), with about half this magnitude seen in those 60-69 years.<br>– Highly significant differences of approximately 1.5-fold were seen per decade across the 60s, 50s and 40s; at least a 2-fold larger rate is apparent between those in their 60s versus <60 years. About a 4-fold association was seen between those aged ≥60 years and 20-49 years.<br>– The associations appeared similar when considering only men or women, although the absolute rates for women were lower across all age groups above 30 years. |
| ICU admission (people with COVID-19) | O'Brien 2020 (a) | – The highest ICU admission rates were in those aged 70-79 years (rates significantly lower in the >80 year age group), with smaller rates, but not by a large amount, in those in their 60s.<br>– People in their 60s appear to have a 1.5 to 2-fold higher rate of ICU admission compared with those in their 50s, and at least a 2-fold larger rate than those <60 years. About a 3-4 fold association was seen between those ≥60 and 20-49 years.<br>– The associations appeared similar when considering only men or women, although the absolute rates for women were lower across all age groups above 30 years. |
| Mortality (people with COVID-19) | Fisman 2020 (a) | – In multivariate regression analysis, adjusted odds of mortality (95% CI) by age (per 10 year increment): 2.42 (1.78, 3.29), p<0.001 |
| | O'Brien 2020 (b) | – Of all closed cases (recovered or died; n = 69,409), the case fatality rate was about 2-4 fold higher I those ≥70 years versus those in their 60s, and at least 2-fold higher for those in their 60s versus 50s. Similar findings were found when considering deaths per 100,000 in the entire Canadian population based on census data. |
| | PHAC COVID -19 SET 2020 | – The mortality rate was less than 1% in all age groups up until the age of 50 years, and then increased rapidly with age, with a rate of 1.2% for those aged 50 to 59 years, 5.9% in those 60-69, and increasing up to 34.4% for those aged 80 years and over. |
| | Wang 2020 | – When excluding long-term care residents and people living in shelters, the relative risk of mortality was about 2-fold higher for those in their 60s versus those 50-59 y, although absolute rates were low (0.7 and 0.3%). |
| **Gender identity/sex** | | |

| Outcome (population) | Study | Key findings |
|---|---|---|
| Hospitalization (people with COVID-19) | O'Brien 2020 (a) | – The rate of hospitalization is lower among females than males in all age categories >30 years. When health care workers, long-term care residents, and school/daycare workers/attendees were excluded, a significant reduction of female cases for hospitalization was only observed in age categories >50 years. |
| ICU admission (people with COVID-19) | O'Brien 2020 (a) | – The rate of ICU admission is lower among females than males in all age categories >30 years. When health care workers, long-term care residents, and school/daycare workers/attendees were excluded, a significant reduction of female cases for ICU admission was only observed in age categories >50 years. |
| Mortality (people with COVID-19) | O'Brien 2020 (a) | – A significantly lower mortality rate was observed in females compared with males within each age category. |
| | PHAC COVID-19 SET 2020 | – Case fatality rate was higher among males (8.5%) than females (7.9%). |
| | Wang 2020 | – In both long-term care and non-long-term care settings, the case fatality proportion appeared to be lower among females (2.8 to 22.8% depending on age group in non-long-term care population) compared to males (7.1 to 30.5%). |
| **Living in long-term care** | | |
| Mortality (general population) | Fisman 2020 (b) | – 229/1,731,315 (<0.1%) individuals >69 years in the general community-dwelling population and 83/79,498 (0.1%) long-term care residents died (IRR (95% CI) 13.1 (9.9, 17.3)); denominators based on estimates and number of facility beds.<br>– Comparisons with other general community-living population ages:<br>○ All adults (n = 14,566,547): 90.4 (68.9, 117.6)<br>○ Aged ≥60 years (n = 3,447,427): 23.1 (17.6, 30.2)<br>○ Aged ≥80 years (n = 642,571): 7.6 (5.5, 10.4)<br>– The median IRR (95% CI) for death among long-term care residents compared with community-living adults rose from 8.03 (1.96, 23.32) on March 29 to 87.28 (6.44, 769.76) by April 11, 2020 |
| Mortality (people with COVID-19) | Fisman 2020 (a) | – Adjusted odds (aOR (95% CI)) of death among cases (n = 1,734) residing in long term care compared with cases residing in the general population in Ontario: 6.24 (2.95, 13.21) |
| | Liu 2020 | – Ontario (long-term care residences): 1,817 deaths (mortality in those with COVID-19, 30.5%)<br>– British Columbia (acute care, long term care and independent living residences): 156 deaths (mortality in those with COVID-19, 33.5%) |
| | O'Brien 2020 (b) | – During the first wave of the pandemic and up to the end of May 2020, long term care facilities and retirement homes accounted for more than 80% of all SARS-CoV-2 deaths in Canada. |
| | PHAC COVID -19 SET 2020 | – Mortality in those with COVID-19 among long-term care residents or those living in seniors' homes compared with those in the general population: 203/758 (26.8%) versus 75/9391 (0.8%).<br>– Mortality in those with COVID-19 by age group among long term care residents compared with those in the general population:<br>○ Aged 0-59 years: 3/33 (9.1%) versus 6/8,122 (0.1%) |

| Outcome (population) | Study | Key findings |
|---|---|---|
| | | ○ Aged 60-79 years: 45/213 (21.1%) versus 34/1,098 (3.1%)<br>– Aged 80+ years: 155/512 (30.3%) versus 35/171 (20.5%) |
| | Wang 2020 | – Adjusted risk (aRR (95%CI) of death among cases (n = 13,122) residing in long term care (918/3368, 27.3%) compared with those residing in the general population (516/12,750, 4.0%) in Greater Toronto area: 1.4 (1.1, 1.8), p=0.02 (adjusted for age and sex)<br>○ Stratified by sex for those aged ≥60 years:<br>▪ Males: RR (95% CI) for 60-70 y 2.4 (1.5 – 4.0), 70-80 y 1.3 (1.0 – 1.7) and ≥80 y 1.2 (1.0 – 1.5)<br>▪ Females: RR (95% CI) for 60-70 y ) 7.6 (4.3 – 13.3), 70-80 y 1.8 (1.2 – 2.7), and ≥80 y 1.2 (1.0 – 1.4) |
| **Homelessness** | | |
| Mortality (people with COVID-19) | Wang 2020 | – Adjusted risk (aRR (95% CI) of death among COVID-19 patients (n = 13,122) residing in a shelter (3/372, 0.8%) compared with those residing in the general population in Greater Toronto area (516/12,750, 4.0%): 0.4 (0.0, 2.5), p=0.5 (adjusted for age and sex) |
| | Fisman 2020 (a) | – Odds (OR (95% CI)) of death among COVID-19 patients (n = 1,734) who were homeless compared with those in the general population (not homeless): 0.43 (0.18, 1.06), p=0.07 |
| **Living on a First Nations reserve** | | |
| Hospitalization (people with COVID-19) | Indigenous Services Canada 2021 | – On First Nations reserves, as of January 6, 2021: 443 hospitalizations in 9,715 cases (4.6%) |
| Mortality (people with COVID-19) | Indigenous Services Canada 2021 | – On First Nations reserves, as of January 6, 2021: 92 deaths in 9,715 cases (0.9%)<br>– vs. https://ipac-canada.org/coronavirus-resources.php Jan 15, 2021: 17,538 deaths in 688,891 cases (2.5%) (all ages)<br>– vs. Public Health Agency of Canada COVID-19 Surveillance and Epidemiology Team 2020 rates in those 0-60 are 0.01% to 1.2% |
| **Occupational exposure: healthcare workers** | | |
| Hospitalization (people with COVID-19) | O'Brien 2020 (a) | – Of all healthcare workers (n = 21,367): 407 (1.9%) were hospitalized and 16,631 (77.8%) were not hospitalized (not reported for 19.5%).<br>– Compared with people having any occupation and non-healthcare occupations: within each decade for those 30-70 years, there appears to be at least a 2-fold higher number of cases hospitalized in those with any occupation or other occupations compared with hospitalizations in healthcare workers; absolute numbers in each decade (in those 30-70 years) were higher for males, but the trends in associations with other occupations were similar between sexes. |
| ICU admission (people with COVID-19) | O'Brien 2020 (a) | – Of all healthcare workers (n = 21,367): 168 (0.8%) were admitted to ICU |

| Outcome (population) | Study | Key findings |
|---|---|---|
| | | – Compared with people having any occupation and non-health are occupations: within each decade for those 30-70 years, there appears to be at least a 2-fold higher number of cases admitted to the ICU in those with any occupation or other occupations compared with healthcare workers; absolute numbers in each decade (in those 30-70 years) were higher for males, but the trends in associations with other occupations were similar between sexes. |
| Mortality (people with COVID-19) | Fisman 2020 (a) | – Odds (OR (95% CI)) of death among cases (n = 1,734) who were healthcare workers compared with those in the general population (not healthcare workers): 0.033 (0.017, 0.063), p<0.001 |
| **Occupation: homeless shelter worker** | | |
| Mortality (people with COVID-19) | Fisman 2020 (a) | – Odds (OR (95% CI)) of death among cases (n = 1,734) who work in a homeless shelter compared with those in the general population (did not work in a homeless shelter): 0.17 (0.024, 1.25), p=0.08 |
| **Race or ethnicity: visible minority groups** | | |
| Mortality (general population) | Subedi 2020 | – Age-standardized mortality rates by % visible minority in neighbourhood, per 100,000 population:<br>○ Less than 1% visible minority: 16.9<br>○ 1 to <10% visible minority: 12.7<br>○ 10 to <25% visible minority: 27.3<br>○ 25+% visible minority: 34.5<br>– In Ontario and Quebec, rates in neighbourhoods with >25% vs. <1% visible minorities were 3-fold; they were 10-fold in British Columbia (although absolute numbers much lower in this province)<br>– Male versus female age-standardized death rate by % visible minority in neighbourhood, per 100,000 population:<br>○ Less than 1% visible minority: 18.3 versus 15.6<br>○ 1 to <10% visible minority: 14.7 versus 11.1<br>○ 10 to <25% visible minority: 34.1 versus 22.7<br>○ 25+% visible minority: 41.0 versus 29.7<br>– Age-standardized mortality rate by % Black, per 100,000 population (Montreal):<br>○ Less than 1% Black: 88.1<br>○ 1 to <10% Black: 105.0<br>○ 10 to <25% Black: 124.7<br>○ 25+% Black: 149.3<br>– Age-standardized mortality rate by % South Asian, per 100,000 population (Toronto):<br>○ <1% South Asian: 26.2<br>○ 1 to <10% South Asian: 24.6<br>○ 10 to <25% South Asian: 29.5<br>– 25+% South Asian: 35.0 |
| **Number of comorbidities** | | |

| Outcome (population) | Study | Key findings |
|---|---|---|
| Mortality (people with COVID-19) | O'Brien 2020 (b) | – Of >9,500 COVID-19 involved deaths between March and July 2020, 90% had at least one other cause, condition, or complication reported on the death certificate.<br>– The proportion of those with at least one other disease or condition reported on the death certificate decreases with age, ranging from 93% for those aged 45 to 64 years, to 89% for those aged 85 and older. |
| | PHAC COVID-19 SET 2020 | – Among patients with COVID-19 for whom data were available on pre-existing conditions (cardiac disease, chronic neurological or neuromuscular disorder, diabetes, immunodeficiency disease/condition, liver disease, malignancy, renal disease, respiratory disease) (n = 6,350):<br>○ Aged 0-59 years: none, 0.1%; 1, 1.5%; 2, 0.0%; 3+, 2.6%<br>○ Aged 60-79 years: none, 3.1%; 1, 3.8%; 2, 12.8%; 3+, 25.5%<br>○ Aged ≥80 years: none, 29.9%; 1, 21.8%; 2, 30.2%; 3+, 37.9%<br>○ All ages: none, 1.9%; 1, 5.8%; 2, 17.0%; 3+, 27.9%<br>– Data for risks based long-term care residency indicate that this is an important variable that may have inflated the findings for pre-existing conditions. |
| **Immune compromise** | | |
| Mortality (people with COVID-19) | Fisman 2020 (a) | – The adjusted odds of mortality (95% CI) among immune compromised was 3.56 (1.12 to 11.35). This finding was adjusted for age, long-term care residency, smoking, renal disease, diabetes, and chronic obstructive pulmonary disease.<br>– In univariate analysis, the odds of mortality (95% CI) among those with malignancy was 6.36 (4.80 to 8.44). |
| | O'Brien 2020 (b) | – Of 9,525 deaths due to COVID-19 in Quebec and Ontario between March and July 2020, 1.8% had cancer as an associated cause. |
| **Pre-existing chronic conditions** | | |
| Mortality (people with COVID-19) | Fisman 2020 (a) | – In multivariate regression analysis, chronic obstructive pulmonary disease (aOR 3.26, 95% CI 1.15-9.26) and diabetes (aOR 2.19, 95% CI 1.08-4.42) were associated with increased mortality. Renal disease may be associated with increased mortality (aOR 2.37, 95% CI 0.97 to 5.77) but the confidence interval was wide.<br>– Multivariate model controlled for age, immune compromise, and long-term care residency (as well as other variables in model, described above) |
| | O'Brien 2020 (b) | – Among deaths from COVID-19, comorbidities listed on the death certificate have included dementia or Alzheimer's disease were listed on the death certificate of 42% of females and 33% of males; other conditions listed included pneumonia (33%), hypertension (15%), ischemic heart disease (13%), respiratory failure (13%), renal failure (12%), diabetes (12%), chronic lower respiratory disease (10%), nervous system disorders excluding Alzheimer's disease (8%), and cancer (8%). |
| **Pregnancy** | | |
| Hospitalization (people with COVID-19) | Money 2021 | – Compared to non-pregnant females of childbearing age, the unadjusted risk ratio (95% CI) for hospitalization was 5.33 (4.51-6.20). |

| Outcome (population) | Study | Key findings |
|---|---|---|
| ICU admission (people with COVID-19) | Money 2021 | − Compared to non-pregnant females of childbearing age, the unadjusted risk ratio (95% CI) for ICU admission was 5.88 (3.80-8.22). |
| **Children and young adults** | | |
| Mortality (people with COVID-19) | O'Brien 2020 (b) | − Healthy young adults (<45 years), adolescents, and children have been least likely to develop severe complications, including death; 100% of deaths from COVID-19 in these age groups as of July 31, 2020 had at least one comorbidity. |
| Hospitalization (people with COVID-19) | Panetta 2020 | − Of 10 infants hospitalized, 2 (20%) had chronic conditions, 5 (50%) were male |
| Severe disease (people with COVID-19) | Panetta 2020 | − Among 27 infants with COVID-19, there was no significant difference in clinical manifestations by age (older vs. younger infants) |

aOR=adjusted odds ratio; CI=confidence interval; COVID-19=novel coronavirus 2019; IRR=incidence rate ratio; OR=odds ratio; PHAC=Public Health Agency of Canada; RR=risk ratio; SET=Surveillance and Epidemiology Team

**Appendix 3.** Characteristics of included studies

**A. Characteristics of studies contributing multivariate-adjusted associations for the main analysis, n=123**

| Author, year;<br>Country (setting);<br>Study design;<br>Funding<br>Study period; follow-up | Enrolled cohort;<br>Study sample;<br>Mean age (SD), years [a]<br>Male, proportion | COVID-19 Diagnosis; Data source | P²ROGRESS risk factors, adjusted for in multivariate regression analysis [b] | Outcomes |
|---|---|---|---|---|
| Adrish, 2020 [1]<br>USA (BronxCare Health System)<br>Retrospective cohort<br>NR<br>9 March to 18 May 2020 (follow-up NR) | Hospitalized with COVID-19<br>N=1,173<br>Median (IQR), smokers: 64.0 (54.0-73.0)<br>Median (IQR), non-smokers: 62.0 (52.0-73.0)<br>61.4% | RT-PCR; Electronic health records | Age, sex, and systemic steroids (as well as LDH levels) | Mortality (in-hospital) |
| Ahlström, 2021 [2]<br>Sweden<br>Retrospective cohort<br>Non-industry<br>6 March to 27 May 2020; 27 May 2020 | People with severe COVID-19<br>N=1,981<br>Median (IQR): 61.0 (52-69)<br>74.0% | RT-PCR; Electronic health records | Age, sex, use of immunosuppressants, ischemic heart disease, non-ischemic heart disease, hypertension, type 1 diabetes, type 2 diabetes, stroke, chronic renal failure, chronic obstructive pulmonary disease, asthma, obesity, systemic inflammatory disease, solid organ transplant, cancer (as well as treatments for COVID-19) | Mortality |
| Alkhouli, 2020 [3]<br>Multi-country (30 countries, 36% USA, 64% NR)<br>Retrospective cohort<br>Non-industry<br>20 January to 20 April 2020 (follow-up NR) | People with COVID-19<br>N=14,712<br>52.8 (17.9)<br>43.4% | Lab-confirmed; Electronic health records (TriNetX multinational research database) | Age, race, obesity, chronic obstructive lung disease, diabetes, heart failure, hypertension, stroke history, and nicotine dependence | Mortality |
| Altschul, 2020 [4]<br>USA (New York City; Montefiore Medical Center)<br>Retrospective cohort<br>NR<br>1 March to 16 April 2020 (follow-up NR) | Hospitalized with COVID-19<br>N=2,355<br>65.3 (15.9)<br>46.5% | RT-PCR; Electronic health records | Age, sex, congestive heart failure (as well as admission labs and clinical signs) | Mortality (in-hospital) |
| Alvarez-Garcia, 2020 [5] | Hospitalized with COVID-19<br>N=6,439<br>63.5 (18.0) | RT-PCR;<br>Electronic health records | Age, sex, race, obesity, hypertension, diabetes, coronary artery disease, atrial fibrillation, chronic kidney disease, chronic obstructive | ICU admission<br>Mechanical ventilation<br>Mortality (in-hospital) |

| Author, year;<br>Country (setting);<br>Study design;<br>Funding<br>Study period; follow-up | Enrolled cohort;<br>Study sample;<br>Mean age (SD), years [a]<br>Male, proportion | COVID-19 Diagnosis; Data source | P²ROGRESS risk factors, adjusted for in multivariate regression analysis [b] | Outcomes |
|---|---|---|---|---|
| USA (New York City; 5 Mount Sinai Healthcare System hospitals)<br>Retrospective cohort<br>NR<br>27 February to 26 June 2020; 18 July 2020 | 55.0% | | pulmonary disease, previous treatment with RAASi | |
| An, 2020 [6]<br>South Korea<br>Prospective cohort<br>NR<br>23 January to 2 April 2020; 16 April 2020 | People with COVID-19<br>N=10,237<br>44.97 (19.79)<br>39.9% | Test positive;<br>Korean National Health Insurance Service database | Age, sex, income level, residence, household type, disability (moderate or severe), chronic lung disease or asthma, diabetes, hyperlipidemia, cancer, cardiovascular disease, cerebrovascular disease, metformin use, cancer, hypertension (as well as presence of symptoms and infection route) | Mortality |
| Anantharaman, 2021 [7]<br>USA (Northern California)<br>Retrospective cohort<br>NR<br>25 February to 8 June 2020; 45 days | People with COVID-19<br>N=4,627<br>74.5% 18-59 years; 13.5% 60-69 years; 11.1% 70+ years<br>48% | Unspecified lab test;<br>Electronic medical records | Cancer history, age, sex, race/ethnicity, BMI, Charlson comorbidity index, hypertension, smoking, diabetes, neighbourhood deprivation index | Hospitalization<br>ICU admission<br>Mechanical ventilation<br>Mortality |
| Anderson, 2020 [8]<br>USA (New York City; NewYork-Presbyterian/ Columbia University Irving Medical Center & Allen Hospital)<br>Retrospective cohort<br>Non-industry<br>10 March to 24 April 2020; 10 June 2020 | Hospitalized with COVID-19<br>People with severe COVID-19<br>N=2,466 (152 mechanically ventilated)<br>Median (IQR): 67.0 (54.0-78.0)<br>58.0% | RT-PCR;<br>Electronic health records (New-York-Presbyterian/Columbia University Irving Medical Center Clinical Data Warehouse; COVID-19 test results from Department of Health records) | Age, sex, race/ethnicity, smoking status, hypertension, diabetes, cancer, asthma, COPD, chronic kidney disease, pulmonary heart disease by BMI (stratified by age and sex) | Severe disease (in-hospital mortality or intubation)<br>Mortality (among mechanically ventilated) |
| Attaway, 2020 [9]<br>USA (Ohio & Florida)<br>Retrospective cohort<br>Non-industry<br>8 March to 13 May 2020 (follow-up NR) | People with COVID-19<br>Hospitalized with COVID-19<br>N=2,527 (705 hospitalized)<br>61.0 (15.5<br>47.8% | RT-PCR;<br>Electronic health records (Cleveland Clinic COVID-19 registry) | Age, sex, BMI, cancer, coronary artery disease, diabetes mellitus, hypertension, immunosuppressive therapy and smoking status | Hospitalization<br>ICU admission (among hospitalized)<br>Mechanical ventilation (among hospitalized)<br>Mortality (in-hospital) |

| Author, year;<br>Country (setting);<br>Study design;<br>Funding<br>Study period; follow-up | Enrolled cohort;<br>Study sample;<br>Mean age (SD), years [a]<br>Male, proportion | COVID-19 Diagnosis; Data source | $P^2$ROGRESS risk factors, adjusted for in multivariate regression analysis [b] | Outcomes |
|---|---|---|---|---|
| Azar, 2020 [10]<br>USA (Northern California)<br>Retrospective cohort<br>NR<br>1 January to 8 April 2020 (follow-up NR) | People with COVID-19<br>N=1,052<br>53.0 (95% CI, 51.8-54.1)<br>49.2% | Lab-confirmed;<br>Electronic health records (Sutter Health) | Age, race/ethnicity, sex, insurance type, median income, homeless status, smoking status, Type 2 diabetes, hypertension, depression, congestive heart failure, cardiovascular disease, cancer, chronic obstructive pulmonary disease, asthma | Hospitalization |
| Bailey 2021 [11]<br>USA (Pennsylvania, Ohio, Colorado, Delaware, Florida, Washington, Missouri)<br>Retrospective cohort<br>Non-industry<br>1 January to 8 September 2020 (follow-up NR) | Children with COVID-19<br>N=5,374<br>Age NR<br>48% | RT-PCR; Electronic health records | Race/ethnicity, age, presence of progressive condition, various types of conditions (endocrine, metabolic, malignancy), history of public insurance | Severe disease (pneumonia, sepsis, or respiratory failure) |
| Barron, 2020 [12]<br>England<br>Retrospective cohort<br>No funding<br>1 March to 11 May 2020;<br>72 day observation period | General population<br>N=61,414,470 (hospitalized NR)<br>78.6 (12.1)<br>61.5% | Antigen testing; National Data Repository (includes Master Patient Index, National Diabetes Audit, Bridges to Health national population segmentation & COVID patient notification system) | Age, sex, ethnicity, deprivation quintile, diabetes status | Mortality (in-hospital) |
| Bennett 2021 [13]<br>Ireland<br>Retrospective cohort<br>Non-industry<br>2 March to 31 July 2020; to 14 September 2020, discharge or death | People with COVID-19<br>Hospitalized with COVID-19<br>N=19,789 (2,811 hospitalized)<br>Age NR<br>43.6% | Nucleic acid testing; surveillance data and death certificates | Age, sex, chronic heart disease, chronic neurological conditions, chronic respiratory disease, chronic kidney disease, chronic liver disease, asthma (requiring medication), immunodeficiency including HIV, diabetes, BMI ≥kg/$m^2$ cancer/malignancy, other comorbidity, residential care facility | Hospitalization<br>ICU admission<br>Mortality |
| Berenguer, 2020 [14]<br>Spain (nationwide)<br>Retrospective cohort<br>Non-industry<br>NR to 17 March 2020; 17 April 2020 | Hospitalized with COVID-19<br>N=4,035<br>Median (IQR): 70.0 (56.0-80.0)<br>61.0% | RT-PCR;<br>Electronic health records | Age, sex, cancer, chronic heart disease, chronic kidney disease stage 4, chronic neurological disorder, chronic pulmonary disease not asthma, dementia, diabetes, hypertension, liver cirrhosis, obesity (as well as admission signs and symptoms, vital signs, lab parameters) | Mortality |

| Author, year;<br>Country (setting);<br>Study design;<br>Funding<br>Study period; follow-up | Enrolled cohort;<br>Study sample;<br>Mean age (SD), years [a]<br>Male, proportion | COVID-19 Diagnosis; Data source | $P^2$ROGRESS risk factors, adjusted for in multivariate regression analysis [b] | Outcomes |
|---|---|---|---|---|
| Bowe, 2020 [15]<br>USA (Veteran Affairs Corporate Data Warehouse)<br>Prospective cohort<br>Non-industry<br>1 February to 23 July 2020; 30 July 2020 | Hospitalized with COVID-19<br>N=5,216 veterans<br>Median (IQR): 70.0 (61.0-76.0)<br>94.0% | RT-PCR;<br>Electronic health records maintained by the Veteran Affairs Corporate Data Warehouse | Age, sex, race, BMI, smoking status, cardiovascular disease, type 2 diabetes, hypertension, immunosuppressants (as well as other medications, lab tests and vital signs) | Acute kidney injury<br>Severe disease (acute kidney injury stage 3, acute kidney injury receiving kidney replacement therapy, and death) |
| Bravi, 2020 [16]<br>Italy (Ferrara and Pescara provinces)<br>Retrospective cohort<br>No funding<br>NR to 2 April or 24 April 2020 (by province); median 24 days | People with COVID-19<br>N=1,603<br>58.0 (20.9)<br>47.3% | RT-PCR; National database of pharmacological treatment linked with electronic health records | Age, sex, type 2 diabetes, hypertension, major cardiovascular diseases (heart failure, myocardial infarction, stroke), cancer, chronic obstructive pulmonary diseases (COPD, bronchitis, asthma, pneumonia, emphysema), renal disease | Hospitalization<br>Severe disease (ICU admission or mortality composite) |
| Carter, 2020 [17]<br>Italy & United Kingdom<br>Prospective cohort<br>No funding<br>27 February to 28 April 2020 (follow-up NR) | Hospitalized with COVID-19<br>N=1,564<br>Median (IQR): 74.0 (61.0-83.0)<br>57.7% | Lab-confirmed (95.1%), clinical diagnosis 64 (4.9%); Electronic and manual health records | Location of infection (community vs. nosocomial), age, sex, smoking status, diabetes, hypertension, coronary artery disease, reduced renal function, and clinical frailty scale (as well as CRP levels) | Hospital length of stay |
| Chan, 2020 [18]<br>USA (New York, Mount Sinai Health System)<br>Retrospective cohort<br>No funding<br>27 February to 30 May 2020; 5 June 2020 | Hospitalized with COVID-19<br>N=3,993<br>Median (IQR): 64.0 (56-78)<br>57.3% | RT-PCR;<br>Electronic health records | Age, sex race, chronic kidney disease, congestive heart failure, diabetes mellitus, hypertension, liver disease, peripheral vascular disease (as well as lab values and vitals) | Acute kidney injury, stage 3 |
| Chhiba, 2020 [19]<br>USA (Chicago and surrounding suburbs)<br>Retrospective cohort<br>Non-industry<br>1 March to 15 April 2020; 30 April 2020 | People with COVID-19<br>N=1,526<br><40 y: 27.1%<br>40-69 y: 55.3%<br>>70 y: 17.6%<br>47.0% | RT-PCR; Electronic health records (Northwestern Medicine Enterprise Data Warehouse) | Age, sex, race/ethnicity, smoking, obesity, CAD, diabetes, HTN, OSA, COPD, allergic rhinitis, rhinosinusitis, immunodeficiency | Hospitalization (with or without ICU admission) |

| Author, year;<br>Country (setting);<br>Study design;<br>Funding<br>Study period; follow-up | Enrolled cohort;<br>Study sample;<br>Mean age (SD), years [a]<br>Male, proportion | COVID-19 Diagnosis; Data source | $P^2$ROGRESS risk factors, adjusted for in multivariate regression analysis [b] | Outcomes |
|---|---|---|---|---|
| Clift, 2020 [20]<br>United Kingdom<br>Retrospective cohort<br>Non-industry<br>24 January to 30 June 2020; 28 days follow-up for mortality | General population<br>N=8,256,158<br>NR<br>NR | RT-PCR; Electronic health records (Qresearch primary care database linked to Public Health England testing results) | Age, sex, ethnicity, BMI, asthma, atrial fibrillation, blood cancer, cerebral palsy, chronic liver disease, COPD, congenital heart disease, coronary disease, dementia, diabetes Type 1, diabetes Type 2, epilepsy, heart failure, rare lung diseases (cystic fibrosis, extrinsic allergic alveolitis), severe mental illness, osteoporotic fracture, peripheral vascular disease, pulmonary hypertension or pulmonary fibrosis, previous stroke, rheumatoid arthritis or SLE, venous thromboembolism and treatments | Hospitalization<br>Mortality |
| Cordtz 2020 [21]<br>Denmark<br>Prospective cohort<br>Non-industry<br>1 March to 12 August 2020; to 12 August 2020 | General population<br>Hospitalized with COVID-19<br>N=4,597,229 (2,674 hospitalized)<br>Median 49.8-71.2 years across groups<br>49.3% | Laboratory-confirmed; Danish national registers | Age, sex, inflammatory rheumatic diseases, cardiovascular disease, lung disease, diabetes mellitus, cancer | Hospitalization<br>Severe disease (mechanical ventilation, acute respiratory distress syndrome, or death) |
| Cummings, 2020 [22]<br>USA (2 hospitals in Northern Manhattan)<br>Prospective cohort<br>Non-industry<br>2 March to 1 April 2020; 14 April 2020 | Hospitalized with COVID-19<br>N=1,150<br>Median (IQR): 62.0 (51.0-72.0)<br>66.0% | RT-PCR; Electronic health records | Age, sex, BMI, hypertension, chronic cardiovascular disease, COPD and/or interstitial lung disease, chronic kidney disease, diabetes (as well as symptom duration and vital signs) | Mortality (in-hospital) |
| Czernichow, 2020 [23]<br>France (Paris)<br>Retrospective cohort<br>Non-industry<br>1 February to 30 April 2020; 30 May 2020 | Hospitalized with COVID-19<br>N=5,795<br>59.8 (13.6)<br>65.4% | PCR; Electronic health records (EDS-COVID database) | Age, sex, BMI, cancer, chronic kidney disease, diabetes, dyslipidemia, heart failure, hypertension, sleep apnea, smoking status (stratified by age) | Mortality (in-hospital) |
| de Azambuja, 2020 [24]<br>Belgium<br>Prospective cohort<br>Non-industry | Hospitalized with COVID-19<br>N=10,486<br>67.8 (17.0)<br>53.2% | PCR 91.6%, 7.6% CT-scan and clinical findings, and 0.8% unknown diagnostic method; Electronic health records (Sciensano - | Age, gender, RAAI use, cardiovascular disease, chronic kidney disease, chronic liver disease, chronic lung disease, chronic neurological disease, cognitive disorder, diabetes, | Severe disease (composite of ICU admission, invasive ventilation use and/or death within 30 days of COVID-19 diagnosis) |

| Author, year;<br>Country (setting);<br>Study design;<br>Funding<br>Study period; follow-up | Enrolled cohort;<br>Study sample;<br>Mean age (SD), years [a]<br>Male, proportion | COVID-19 Diagnosis; Data source | P$^2$ROGRESS risk factors, adjusted for in multivariate regression analysis [b] | Outcomes |
|---|---|---|---|---|
| 15 February to 24 May 2020; followed 30 days for mortality | | Belgian public health surveillance database) | haematological cancer, hypertension, immunosuppression including HIV | Mechanical ventilation<br>Mortality (in-hospital) |
| Dennis, 2020 [25]<br>United Kingdom<br>Retrospective cohort<br>Non-industry<br>1 March to 27 July 2020; followed up to 30 days | Hospitalized with COVID-19 and requiring HDU or ICU admission<br>N=19,256<br>67.0 (16.9)<br>60.1% | RT-PCR (91% with swab positive result); Electronic health records | Age, sex, ethnicity, obesity, comorbidities | Mortality (in-hospital) |
| Denova-Gutiérrez [26]<br>Mexico<br>Retrospective cohort<br>No funding<br>27 February to 10 April 2020 (follow-up NR) | People with COVID-19<br>N=3,844<br>45.4 (15.8)<br>58.0% | RT-PCR; Electronic health records (National Epidemiological Surveillance System) | Age, sex, smoking status, history of chronic diseases (cardiovascular disease, chronic kidney disease, immunosuppression), place of care, respiratory medical unit, and drug treatment, hypertension, obesity | Severe disease (pneumonia and other organ failure requiring treatment in ICU) |
| Docherty, 2020 [27]<br>United Kingdom<br>Prospective cohort<br>Non-industry<br>6 February to 19 April 2020; minimum 2 week follow-up | Hospitalized with COVID-19<br>N=21,033<br>Median (IQR): 73.0 (58.0-82.0)<br>59.9% | RT-PCR; routine health records | Age, sex, chronic cardiac disease, chronic pulmonary disease, diabetes, obesity, chronic neurological disorder, dementia, malignancy, moderate/severe liver disease | ICU admission<br>Mortality |
| D'Silva 2020 [28]<br>USA (nationwide)<br>Retrospective cohort<br>Non-industry<br>20 January to 15 August 2020; 30 days | People with COVID-19<br>N=4,748<br>58 (16)<br>21% | RT-PCR; Electronic health records (TriNetX) | Systemic autoimmune rheumatic diseases, age, sex, BMI, race/ethnicity, comorbidities, prior hospitalization | Hospitalization<br>ICU admission<br>Mechanical ventilation<br>Acute renal failure<br>Ischemic stroke<br>Severe disease (composite ICU admission, mechanical ventilation, death)<br>Mortality |
| Ellington, 2020 [29]<br>USA (nationwide)<br>Prospective cohort<br>NR | Women with COVID-19<br>N=91,412<br>Pregnant:<br>25-34 y: 54.4% | Lab-confirmed or antigen test; CDC database from national COVID-19 surveillance | Age, ethnicity, underlying conditions | Hospitalization |

| Author, year;<br>Country (setting);<br>Study design;<br>Funding<br>Study period; follow-up | Enrolled cohort;<br>Study sample;<br>Mean age (SD), years [a]<br>Male, proportion | COVID-19 Diagnosis; Data source | $P^2$ROGRESS risk factors, adjusted for in multivariate regression analysis [b] | Outcomes |
|---|---|---|---|---|
| 22 January to 7 June 2020<br>(follow-up NR) | 35-44 y: 22.1%<br>Non-pregnant:<br>25-34 y: 38.2%<br>35-44 y: 38.3%<br>0% (all female) | | | |
| Esme, 2020 [30]<br>Turkey<br>Retrospective cohort<br>NR<br>11 March to 27 May 2020<br>(follow-up NR) | Hospitalized with COVID-19<br>N=16,942<br>60-64 y: 27.0%<br>65-69 y: 22.0%<br>70-74 y: 19.0%<br>75-79 y: 13.0%<br>≥80 y: 19.0%<br>49.0% | RT-PCR; Turkish Ministry of Health database | Sex, hypertension, diabetes, heart failure, chronic kidney disease. Dementia, cancer (stratified by age) | Mortality (in-hospital) |
| Floyd 2021 [31]<br>USA (Philadelphia, PA)<br>Retrospective cohort<br>Non-industry<br>17 March to 26 August 2020 (follow-up NR | Children with COVID-19<br>N=979<br>28% 0-4 years, 23% 5-11 years, 34% 12-17 years, 15% 18-21 years<br>51% | RT-PCT; Electronic health recrods | Current asthma (based on EHR registry definition; diagnosis or treatment in past year, persistent asthma); age, sex, race, ethnicity, obesity, number of chronic conditions | Hospitalization |
| Fond, 2020 [32]<br>France (nationwide)<br>Retrospective cohort<br><br>NR<br>1 February 2020 to 9 June 2020 (follow-up NR) | Hospitalized with COVID-19<br>N=50,750<br>Median (IQR): 71.0 (57.0-83.0)<br>56.8% | ICD-10 codes for lab-confirmed COVID;<br>French national hospital database | Age, sex, social deprivation, schizophrenia, smoking status, overweight and obesity, Charlson Comorbidity Index score (stratified by age) | ICU admission<br>Mortality (in-hospital) |
| Fresan, 2020 [33]<br>Spain (Navarra region)<br>Prospective cohort<br>Non-industry<br>1 March to 30 April 2020; followed 30 days for mortality | People with COVID-19<br>Hospitalized with COVID-19<br>N=433,995 (1,105 hospitalized)<br>NR<br>50.0% | RT-PCR and rapid antibody testing;<br>Regional health service electronic records | Age, sex, country of origin, municipality size, annual taxable income level, primary health care visits in prior 12 months, hospitalization in prior 12 months, smoking status, hypertension, and major chronic conditions (stratified by age, and sex) | Hospitalization<br>Severe disease (ICU admission or mortality composite among hospitalized) |
| Garazzino, 2021 [34]<br>Italy (nationwide)<br>Retrospective cohort | Children with COVID-19<br>N=759<br>Mean (IQR): 7.3 (1.4-12.4) | RT-PCR or antibody tests; health records | Age, sex, chronic comorbidities, immunosuppression (as well as more than one infected family member, symptoms) | Hospitalization<br>Mechanical ventilation<br>ICU admission |

| Author, year;<br>Country (setting);<br>Study design;<br>Funding<br>Study period; follow-up | Enrolled cohort;<br>Study sample;<br>Mean age (SD), years [a]<br>Male, proportion | COVID-19 Diagnosis; Data source | $P^2$ROGRESS risk factors, adjusted for in multivariate regression analysis [b] | Outcomes |
|---|---|---|---|---|
| NR<br>Up to 15 September 2020; at least 2 weeks | 56.1% | | | Severe disease (pneumonia, severe acute respiratory illness, acute respiratory distress syndrome, neurological disturbances, severe dehydration requiring intravenous rehydration, severe bacterial supra-infection, specific involvement of a single organ/apparatus requiring hospitalization, multisystem inflammatory syndrome) |
| Geretti, 2020 [35]<br>United Kingdom (England, Scotland, Wales)<br>Prospective cohort<br>Non-industry<br>17 January to 18 June 2020; followed 28 days for mortality | Hospitalized with COVID-19<br>N=47,592<br>Median (IQR)<br>HIV +ve: 56.0 (49.0-62.0)<br>HIV -ve: 74.0 (60.0-84.0)<br>57.3% | RT-PCR (90.5%); Electronic health records (ISARIC WHO CCP-UK) | Age, sex, ethnicity, obesity, chronic cardiac disease, chronic haematological disease, chronic neurological disease, chronic pulmonary disease, chronic renal disease, dementia, diabetes, liver disease & malignancy (as well as hospital admission date & indeterminate/probable hospital acquisition of COVID-19) | ICU admission<br>Mortality |
| Giorgi Rossi, 2020 [36]<br>Italy (Reggio Emilia province)<br>Prospective cohort<br>Non-industry<br>27 February to 2 April 2020; 3 April 2020 | People with COVID-19<br>N=2,653<br><51 y: 696<br>51-60 y: 528<br>61-70 y: 413<br>71-80 y: 420<br>≥81 y: 596<br>50.1% | PCR; SARS-CoV-2 special database (Province of Reggio Emilia, Italy) and electronic health records | Age, sex, Charlson Index, place of residence (as well as calendar period & time from symptom to diagnosis) | Hospitalization<br>Mortality |
| Gottlieb, 2020 [37]<br>USA (Rush University Medical Center, Chicago)<br>Retrospective cohort<br>NR<br>4 March to 21 June 2020 (follow-up NR) | People with COVID-19<br>Hospitalized with COVID-19<br>N=8,673 (1,483 hospitalized)<br>Median (IQR): 41.0 (29.0-54.0)<br><br>46.6% | Lab-confirmed;<br>Electronic health records | Age, sex, race, ethnicity, tobacco use, asthma, COPD, hypertension, hyperlipidemia, diabetes, prior cerebrovascular event (stroke), coronary artery disease, congestive heart failure, chronic kidney disease, current end-stage renal disease, cirrhosis, obstructive sleep apnea, bloodborne | Hospitalization<br>ICU admission (among hospitalized) |

| Author, year;<br>Country (setting);<br>Study design;<br>Funding<br>Study period; follow-up | Enrolled cohort;<br>Study sample;<br>Mean age (SD), years [a]<br>Male, proportion | COVID-19 Diagnosis; Data source | P$^2$ROGRESS risk factors, adjusted for in multivariate regression analysis [b] | Outcomes |
|---|---|---|---|---|
| | | | cancer, solid organ cancer, HIV, solid organ transplant, BMI | |
| Gotzinger, 2020 [38]<br>Europe (21 countries, 77 healthcare institutions)<br>Retrospective cohort<br>Non-industry<br>1 April to 24 April 2020 (follow-up NR) | People with COVID-19<br>N=582<br>Median (IQR): 5.0 (0.5-12.0)<br>53.0% | RT-PCR; Electronic health records (Pediatric Tuberculosis Network European Trials Group) | Age, sex, pre-existing medical conditions (as well as signs or symptoms) | ICU admission |
| Grasselli, 2020 [39]<br>Italy (Lombardy)<br>Retrospective cohort<br>Non-industry<br>20 February to 22 April 2020; 30 May 2020 | People with severe COVID-19)<br>N=3,988<br>Median (IQR): 63.0 (56.0-69.0)<br>80.0% | RT-PCR; Regional Health System Database | Age, sex, pre-existing comorbidities, (as well as clinical findings & medications) | Mortality (in-hospital & in-ICU) |
| Guerrero-Torres, 2020 [40]<br>Mexico<br>Retrospective cohort<br>NR<br>27 February to 31 August (follow-up NR) | Hospitalized with COVID-19<br>N=25,771<br>57.4 (15.7)<br>63.2% | PCR; Electronic health records (National Epidemiological Surveillance System) | Age, sex, comorbidities, smoking status, state of residence, type of care institution (as well as time from onset of symptoms to first evaluation) | Mortality |
| Gupta, 2020 (a) [41]<br>USA (65 hospitals across the country)<br>Prospective cohort<br>NR<br>4 March to 4 April 2020; median follow-up 16 days (IQR 8-28 days) | People with severe COVID-19<br>N=2,215<br>60.5 (14.5)<br>64.8% | Lab-confirmed; Electronic health records | Age, sex, race, hypertension, diabetes, BMI, coronary artery disease, congestive heart failure, chronic obstructive pulmonary disease, current smoking status, active cancer (as well as lab values assessed at ICU admission) | Mortality<br>Mortality (among mechanically ventilated on day 1 of ICU admission) |
| Gupta, 2020 (b) [42]<br>USA<br>Prospective cohort<br>No funding<br>4 March to 11 April 2020; 1 August 2020 | People with severe COVID-19<br>N=3,099 (637 with acute kidney injury)<br>Median (IQR): 62.0 (51.0-71.0)<br>64.6% | Lab-confirmed;<br>Electronic health records (STOP-COVID database) | Age, sex, race, comorbidities, (as well as days from hospital to ICU admission, illness severity on admission, lab values, secondary infection on day 1, altered mental status on day 1, shock, hospital size, regional density of COVID-19-quartiles by county) | Acute kidney injury<br>Mortality (among ICU admitted and with acute kidney injury) |

| Author, year;<br>Country (setting);<br>Study design;<br>Funding<br>Study period; follow-up | Enrolled cohort;<br>Study sample;<br>Mean age (SD), years [a]<br>Male, proportion | COVID-19 Diagnosis; Data source | $P^2$ROGRESS risk factors, adjusted for in multivariate regression analysis [b] | Outcomes |
|---|---|---|---|---|
| Hajifathalian, 2020 [43]<br><br>USA (1 hospital in Manhattan, New York City)<br>Retrospective cohort<br>NR<br>4 March to 9 April 2020 (follow-up NR) | People with COVID-19<br>N=1,059 (768 hospitalized)<br><br>61.1 (18.3)<br>58.0% | RT-PCR; Medical records | Age, sex, race/ ethnicity, BMI, pre-existing comorbidities (as well as vital signs) | Severe disease (ICU admission/mortality composite) |
| Hamer, 2020 (a) [44]<br>England (UK Biobank)<br>Prospective cohort<br>Non-industry<br>16 March to 26 April 2020; 26 April 2020 | General population<br>N=387,109<br>56.2 (8.0)<br>44.9% | RT-PCR;<br>UK-Biobank, includes self-report of risk factors | Age, sex, education, ethnicity, diabetes, hypertension, cardiovascular disease (heart attack, angina, or stroke), lifestyle score, smoking, physical activity, alcohol consumption, BMI category | Hospitalization |
| Hamer, 2020 (b) [45]<br>England (UK Biobank)<br>Retrospective cohort<br>No funding<br>16 March to 26 April 2020 (follow-up NR) | General population<br>N=334,329<br>56.4 (8.1)<br>45.5% | RT-PCR;<br>UK Biobank Study (clinic visit; self-report); Public Health England | Age, sex, smoking status, physical activity, alcohol use, ethnicity, education, diabetes, cardiovascular disease (heart attack, angina, or stroke), hypertension, BMI | Hospitalization |
| Harrison, 2020 [46]<br>USA<br>Retrospective cohort<br>No funding<br>20 January to 26 May 2020; median follow-up 54 days (IQR 36-68) | People with COVID-19<br>N=31,461<br>Median (IQR): 50.0 (35.0-63.0)<br>45.5% | Lab-confirmed plus ICD-10 codes; Electronic health records (TriNetX network) | Age, sex, ethnicity, myocardial infarction, congestive heart failure, peripheral vascular disease, cerebrovascular disease, dementia, chronic pulmonary disease, rheumatic disease, peptic ulcer disease, liver disease (mild, moderate, severe), diabetes, hemiplegia/paraplegia, renal disease, any malignancy, metastatic solid tumor, AIDS/HIV | Mortality |
| Hernandez-Vasquez, 2020 [47]<br>Mexico<br>Retrospective cohort<br>No funding<br>Through 18 May 2020 (follow-up NR) | Patients with COVID-19<br>N=51,053<br>46.6 (15.8)<br>57.6% | RT-PCR;<br>Secretaría de Salud de México (national epidemiologic surveillance database) | Age, sex, smoking status (stratified by sex) | Mortality |

| Author, year;<br>Country (setting);<br>Study design;<br>Funding<br>Study period; follow-up | Enrolled cohort;<br>Study sample;<br>Mean age (SD), years [a]<br>Male, proportion | COVID-19 Diagnosis; Data source | $P^2$ROGRESS risk factors, adjusted for in multivariate regression analysis [b] | Outcomes |
|---|---|---|---|---|
| Hirsch, 2020 [48]<br>USA (New York City, Northwell health)<br>Retrospective cohort<br>NR<br><br>1 March to 5 April 2020 (follow-up NR) | Hospitalized with COVID-19<br>N=5,449<br>Median (IQR): 64.0 (52.0-75.0)<br><br>60.9% | RT-PCR; Heath records (managed by Sunrise Clinical Manager) | Age, sex, race, diabetes, hypertension, cardiovascular disease, obesity, HIV, cancer, ACE-I or ARB use (as well as tertiary hospital, mechanical ventilation and medications) | Acute kidney injury |
| Huh, 2020 [49]<br>South Korea<br>Retrospective cohort<br>Non-industry<br>NR | People with COVID-19<br>N=2,231<br>7.3% 20-29, 10.4% 30-39, 17.1% 40-49, 30.3% 50-59, 22.3% 60-69, 9.5% 70-79, 3.0% ≥80 | RT-PCR; Electronic health records (National Health Insurance Service database, and Korea Centers for Disease Control and Prevention COVID-19 registry) | Age, sex, Charlson comorbidity index, BMI, asthma, cancer, chronic heart disease, chronic kidney disease, chronic liver disease, chronic lung disease, diabetes, hypertension, rheumatologic disease, chronic rheumatologic disease, coverage for low income | Severe disease (supplementary oxygen, high-flow nasal cannula, non-invasive ventilation, mechanical ventilation, extracorporeal membrane oxygenation, or mortality) |
| Iaccarino, 2020 [50]<br>Italy (nationwide)<br>Retrospective cohort<br>Non-industry<br>9 March to 9 April 2020 (follow-up NR) | Hospitalized with COVID-19<br>N=1,591<br>66.5 (0.4)<br>64.0% | RT-PCR; Hospital charts | Age, sex, comorbidities (as well as medications) | Mortality (in-hospital) |
| Ioannou, 2020 [51]<br>USA<br>Prospective cohort<br>Non-industry<br>28 February to 14 May; 22 June 2020 | People with COVID-19<br>N=10,131<br>63.6 (16.2)<br>91.0% | RT-PCR; US Veterans Health Administration database | Age, sex, race, ethnicity, location (urban vs. rural), Charlson comorbidity index, asthma, BMI, cancer, cerebrovascular disease, chronic kidney disease, COPD, cirrhosis, coronary heart failure, diabetes, dialysis, hyperlipidemia, hypertension, obesity hypoventilation, obstructive sleep apnea, alcohol dependence, smoking status (as well as COVID-19 related deaths per million residents) | Hospitalization<br>ICU admission<br>Mortality |
| Izurieta, 2021 [52]<br>USA (Medicare beneficiaries)<br>Retrospective cohort<br>Non-industry | General population (all over 65 years)<br>N=2,533,329<br>Median 73.0<br>44.4% | RT-PCR; Electronic medical records | Chronic conditions, social factors (deprivation index), frailty, immunocompromise (immunocompromising conditions or use of immunosuppressive drugs) (as well as influenza vaccination) | Hospitalization<br>Mortality |

| Author, year;<br>Country (setting);<br>Study design;<br>Funding<br>Study period; follow-up | Enrolled cohort;<br>Study sample;<br>Mean age (SD), years [a]<br>Male, proportion | COVID-19 Diagnosis; Data source | P²ROGRESS risk factors, adjusted for in multivariate regression analysis [b] | Outcomes |
|---|---|---|---|---|
| 1 April 2020 to 8 May 2020; at least 21 days | | | | |
| Izzy, 2020 [53]<br>USA (Massachusetts)<br>Prospective cohort<br>NR<br>1 February to 14 April 2020; 25 April 2020 | People with COVID-19<br>N=5,190 (1,489 hospitalized)<br>Median (IQR): 52.0 (36.0-66.0)<br>46.0% | Nucleic acid testing; Electronic health records maintained by Mass General Brigham | Age, sex, smoking status, BMI, diabetes, hyperlipidemia, hypertension, obstructive lung disease, interstitial lung disease, coronary artery disease, congestive heart failure, cerebrovascular disease, obstructive sleep apnea, chronic kidney disease, transplantation, autoimmune disease, malignancy (stratified by ethnicity, and sensitivity analysis with median household income) | Hospitalization<br>ICU admission (among hospitalized) |
| Jakob, 2020 [54]<br>Europe (112 LEOSS partner sites, 98.1% Germany)<br>Retrospective cohort<br>No funding<br>16 March to 14 May 2020 (follow-up NR) | People with COVID-19 (92.7% recruited from hospitals)<br>N=2,155<br>≤14 years: 1.2%<br>15-25 years: 2.7%<br>26-45 years: 14.7%<br>46-65 years: 34.4%<br>66-85 years: 39.6%<br>>85 years: 7.4%<br>59.7% | RT-PCR; data from the Lean European Open Survey on SARS-CoV-2-Infected Patients (LEOSS) cohort study | Age, sex, cardiovascular disease, diabetes, pulmonary disease | Severe disease (oxygen supplementation, mechanical ventilation, clinically meaningful increase of prior oxygen home therapy, new cardiac arrhythmia, new pericardial effusion >1cm, new heart failure with pulmonary edema, congestive hepatopathy or peripheral edema, need for catecholamines, qSOFA ≥2, acute renal failure in need of dialysis, liver failure with Quick < 50%, composite) |
| Ji, 2020 [55]<br>South Korea<br>Retrospective cohort<br>Non-industry<br>NR to 15 May 2020 (follow-up NR) | People with COVID-19<br>N=7,341<br>47.1 (19.0)<br>40.5% | RT-PCR; HIRA insurance claims database and Korean Center for Disease Control database | Age, sex, residence, Charlson comorbidity index, healthcare utilization, comorbidities | Severe disease (oxygen therapy, mechanical ventilation, extracorporeal membrane oxygenation, and cardiopulmonary resuscitation) |
| Jiminez, 2020 [56]<br>Spain (Madrid)<br>Retrospective cohort<br>No funding | Hospitalized with COVID-19<br>N=1,549<br>Median (IQR): 69.0 55.0-81.0 | RT-PCR; Electronic health records | Age, sex, migrant status, cardiovascular disease, hypertension, diabetes, smoking status, COPD, OSAS, neurological disease, chronic kidney | Mortality (in-hospital) |

| Author, year;<br>Country (setting);<br>Study design;<br>Funding<br>Study period; follow-up | Enrolled cohort;<br>Study sample;<br>Mean age (SD), years [a]<br>Male, proportion | COVID-19 Diagnosis; Data source | $P^2$ROGRESS risk factors, adjusted for in multivariate regression analysis [b] | Outcomes |
|---|---|---|---|---|
| 1 March to 28 May 2020;<br>28 May 2020 | 57.5% | | disease, cancer (as well as biomarkers & clinical features on presentation) | |
| Kabarriti, 2020 [57]<br>USA (New York City)<br>Retrospective cohort<br>NR<br>14 March to 15 April 2020;<br>27 April 2020 | People with COVID-19<br>N=5,902<br>Median (IQR): 58.0 (44.0-71.0)<br>46.0% | RT-PCR; Electronic health records (Bronx Montefiore Health System) | Age, sex, race/ethnicity, SES, BMI, cancer, cardiovascular disease, chronic pulmonary disease, dementia, diabetes, hemiplegia or paraplegia, HIV/AIDS, hypertension, kidney disease, liver disease, peptic ulcer | Mortality |
| Kaeuffer, 2020 [58]<br>France (Strasbourg and Mulhouse)<br>Prospective cohort<br>Non-industry<br>March 2020; up to day 7 following hospitalization | Hospitalized with COVID-19<br>N=1,045<br>66.3 (16.0)<br>58.6% | RT-PCR; Electronic medical records | Age, sex, body mass index, hypertension, diabetes, chronic lung disease, immunosuppression, chronic kidney disease (as well as symptoms, biological findings) | Severe disease (ICU admission or mortality)<br>Mortality |
| Kim, 2020 (a) [59]<br>USA (154 acute-care hospitals in 74 counties in 13 states)<br>Retrospective cohort<br>Non-industry<br>1 March to 2 May 2020 (follow-up NR) | Hospitalized with COVID-19<br>N=2,491<br>Median (IQR): 62.0 (50.0-75.0)<br>53.2% | Lab-confirmed;<br>Laboratory and reportable condition databases, hospital infection control databases, electronic medical records, and/or reviews of hospital discharge records | Age, sex, race and ethnicity, current or former smoker; a history of hypertension, obesity, diabetes, chronic lung disease, cardiovascular disease, neurologic disorders, renal disease, immunosuppression, hematologic conditions, rheumatologic/autoimmune conditions (as well as angiotensin receptor blocker use prior to hospitalization) | ICU admission<br>Mortality (in-hospital) |
| Kim, 2020 (b) [60]<br>USA (New York City, Northwell health system)<br>Retrospective cohort<br>Non-industry<br>1 March to 27 April 2020;<br>12 May 2020 | Hospitalized with COVID-19<br>N=10,861<br>Median (IQR): 65.0 (54.0-77.0)<br>59.6% | PCR; Electronic health records (Northwell Health System) | Age, sex, race/ethnicity, BMI, asthma, cancer, chronic kidney disease, COPD, coronary artery disease, diabetes mellitus, end stage renal disease, hypertension, smoking status, hospital type | Mechanical ventilation<br>Mortality (in-hospital) |
| King, 2020 [61]<br>USA<br>Prospective cohort<br>Non-industry<br>2 March to 18 July 2020; 19 August 2020 | People with COVID-19<br>N=3,681<br>Median (IQR): 64.8 (53.7-73.4)<br>92.6% | Lab-confirmed;<br>US Veterans Health Administration database | Age, sex, race/ethnicity, Charlson comorbidity index, AIDS, asthma, cancer, metastatic cancer, cerebrovascular accident, chronic pulmonary disease, dementia, diabetes (without or with complications), hypertension, liver disease (mild, severe), myocardial infarction, peptic | Mortality |

| Author, year;<br>Country (setting);<br>Study design;<br>Funding<br>Study period; follow-up | Enrolled cohort;<br>Study sample;<br>Mean age (SD), years [a]<br>Male, proportion | COVID-19 Diagnosis; Data source | $P^2$ROGRESS risk factors, adjusted for in multivariate regression analysis [b] | Outcomes |
|---|---|---|---|---|
| | | | ulcer disease, peripheral vascular disease, plegia, renal disease, rheumatological disease | |
| Kjeldsen, 2021 [62]<br>Denmark<br>Retrospective cohort<br>Industry and non-industry<br>1 March to 31 October 2020 (follow-up NR) | Hospitalized with COVID-19<br>N=2,943<br>Median (IQR):<br>Exposed 74.0 (63-80)<br>Controls 69.0 (54-80)<br>55.6% | RT-PCR; Electronic medical records | Age, sex, Charlson comorbidity index, chronic inflammatory diseases | Mechanical ventilation<br>Mortality |
| Klang, 2020 [63]<br>USA (New York City, 5 Mount Sinai hospital campuses)<br>Retrospective cohort<br>1 March to 17 May 2020 (follow-up NR) | Hospitalized with COVID-19<br>N=3,406<br>Median age (IQR)<br>Survivors<br>Age >50 y: 68.0 (60.0-77.0)<br>Age ≤50 y: 40.0 (34.0-46.0)<br>Non-survivors<br>Age >50 y: 76.0 (67.0-84.0)<br>Age ≤50 y: 46.5 (42.8-49.0)<br>57.6% | PCR; Electronic health records (5 campus hospitals in New York City) | Age, sex, coronary artery disease, congestive heart failure, hypertension, diabetes, hyperlipidemia, chronic kidney disease, cancer, smoking, BMI, race (stratified by age) | Mechanical ventilation<br>Mortality (in-hospital) |
| Ko, 2020 [64]<br>USA<br>Prospective cohort<br>Non-industry<br>1 March to 23 June 2020; 23 June 2020 | General population<br>N=5,416<br>Median (IQR): 55.0 (42.0-67.0)<br>53.0% | Lab-confirmed; COVID-19–Associated Hospitalization Surveillance Network (COVID-NET) | Age, sex, race/ethnicity | Hospitalization |
| Kohl, 2020 [65]<br>United Kingdom (2 hospitals, Derby)<br>Retrospective cohort<br>No funding<br>5 March to 12 May 2020 (follow-up NR) | Hospitalized with COVID-19<br>N=1,161<br>72.1 (16.1)<br>56.6% | RT-PCR; Electronic health records | Age, sex, ethnicity, cancer, cerebrovascular disease, chronic kidney disease, chronic liver disease, chronic lung disease, congestive cardiac failure, dementia, diabetes with complications, myocardial infarction, paraplegia, peripheral vascular disease, care home residence (as well as treatments) | Acute kidney injury<br>Mortality (in-hospital) |
| Kragholm, 2020 [66]<br>Denmark (nationwide)<br>Retrospective cohort | People with COVID-19<br>N=4,842<br>Median (IQR)<br>Males: 57.0 (42.0-73.0)<br>Females: 52.0 (38.0-71.0) | RT-PCR; Danish National Patient Registry, Danish Civil Registration System and Danish Prescription Registry | Age, alcohol use, obesity, hypertension, diabetes, chronic obstructive pulmonary disease, sleep apnea, prior myocardial infarction, chronic ischemic heart disease, heart failure, atrial fibrillation or flutter, stroke, | Severe disease (severe COVID-19, ICU admission or mortality composite)<br>ICU admission<br>Mortality |

| Author, year;<br>Country (setting);<br>Study design;<br>Funding<br>Study period; follow-up | Enrolled cohort;<br>Study sample;<br>Mean age (SD), years [a]<br>Male, proportion | COVID-19 Diagnosis; Data source | P$^2$ROGRESS risk factors, adjusted for in multivariate regression analysis [b] | Outcomes |
|---|---|---|---|---|
| End of February to 16 May 2020; followed up to 30 days | 47.1% | | peripheral artery disease, liver disease, rheumatic disease, chronic kidney disease, cancer (stratified by sex) | |
| Kummer, 2020 [67]<br>USA (New York City, 5 Mount Sinai hospital campuses)<br>Retrospective cohort<br>No funding<br>1 March to 1 May 2020 (follow-up NR) | Hospitalized with COVID-19<br>N=3,248<br>Median (IQR)<br>History of stroke: 75.0 (65.0-83.0)<br>No history of stroke: 66.0 (55.0-77.0)<br>58.2% | PCR; Electronic health records (5 hospital campuses across Mount Sinai health system) | Age, sex, hypertension, coronary artery disease, diabetes, dyslipidemia, congestive heart failure, atrial fibrillation, chronic kidney disease, obesity, COPD, asthma, active smoking, malignancy, stroke | Mortality (in-hospital) |
| Kundi, 2020 [68]<br>Turkey (nationwide)<br>Retrospective cohort<br>Non-industry<br>11 March to 22 June 2020; 20 July 2020 | Hospitalized with COVID-19<br>N=18,234<br>74.1 (7.4)<br>46.6% | RT-PCR; Electronic health records (e-Pulse and National Healthcare Information System of Turkey) | Age, sex, coronary artery disease, coronary artery bypass graft, congestive heart failure, valvular heart disease, hypertension, peripheral vascular disease, cerebrovascular disease, COPD, diabetes, liver disease, renal failure, iron deficiency anemia, rheumatoid disease, peptic ulcer disease, depression, cancer, substance abuse, alcohol abuse, acquired immunodeficiency syndrome, hospital frailty risk score | Hospital stay (>10 days)<br>ICU admission<br>Mechanical ventilation<br>Mortality |
| Lassale, 2020 [69]<br>United Kingdom<br>Retrospective cohort<br>No funding<br>16 March to 26 April 2020 (follow-up NR) | People with COVID-19<br>N=340,966<br>56.2 (NR)<br>46.0% | RT-PCR; UK Biobank | Age, sex, ethnicity, education, townsend score, physical activity, smoking, alcohol use, BMI, household size, hypertension, cardiovascular disease, chronic bronchitis, ever seen psychiatrist (as well as lab values) | Hospitalization |
| Lee, 2020 (a) [70]<br>South Korea<br>Retrospective cohort<br>Non-industry<br>1 January to 15 May 2020 (follow-up NR) | People with COVID-19<br>N=2,640<br>Mental illness: 57.8 (16.8)<br>No mental illness: 58.3 (16.6)<br>39.1% | RT-PCR; South Korean national health insurance claims database | Mental illness, age, sex, region of residence, diabetes, cardiovascular disease, cerebrovascular disease, COPD, asthma, hypertension, chronic kidney disease, Charlson comorbidity index | Severe disease (ICU admission, mechanical ventilation or mortality composite)<br>Mortality |

| Author, year;<br>Country (setting);<br>Study design;<br>Funding<br>Study period; follow-up | Enrolled cohort;<br>Study sample;<br>Mean age (SD), years [a]<br>Male, proportion | COVID-19 Diagnosis; Data source | $P^2$ROGRESS risk factors, adjusted for in multivariate regression analysis [b] | Outcomes |
|---|---|---|---|---|
| Lee, 2020 (b) [71]<br>South Korea<br>Retrospective cohort<br>Non-industry<br>NR to 15 May 2020; 15 May 2020 | People with COVID-19<br>N=7,339<br>47.1 (19.0)<br>40.1% | RT-PCR; Korean Health Insurance Claim Data (98% of population) | Age, sex, location, influenza, tuberculosis, COPD, pneumonia, asthma, diabetes, chronic kidney disease, liver disease, hypertension, cardiovascular disease, malignancy, HIV (as well as treatments and medications) | Severe disease (oxygen therapy, mechanical ventilation, CPR, or extracorporeal membrane oxygenation)<br>Mortality |
| Lee, 2020 (c) [72]<br>South Korea (Daegu)<br>Retrospective cohort<br>Non-industry<br>NR to 8 March 2020<br>(follow-up NR) | People with COVID-19<br>People with severe COVID-19<br>N=4,742<br>41.8 (19.0)<br>45.5% | RT-PCR; Health Insurance Review & Assessment Service database | Age, sex, insurance type, facility location, malignancy, COPD, ischemic heart disease, hypertension, diabetes | ICU admission<br>Mechanical ventilation (among ICU admitted) |
| Leon-Abarca, 2020 [73]<br>Mexico<br>Retrospective cohort<br>NR<br>NR | General population (children)<br>N=21,161<br>NR<br>NR | RT-PCR; Electronic health records (Mexican Open Registry) | Age, sex, area of residence, immunodeficiencies, asthma, obesity, cardiovascular diseases, chronic kidney disease, hypertension, diabetes | Hospitalization<br>ICU admission<br>Mortality |
| Loffi, 2020 [74]<br>Italy (Lombardy)<br>Retrospective cohort<br>No funding<br>21 February to 31 March 2020; 4 May 2020 | Hospitalized with COVID-19<br>N=1,252<br>64.7 (15.5)<br>63.7% | RT-PCR; Electronic health records; telephone follow up for discharged patients | Coronary artery disease, age, sex, smoking status, hypertension, hyperlipidemia, diabetes, chronic kidney disease, prior cerebrovascular event, atrial fibrillation (as well as left ventricular ejection fraction <35%) | Mortality |
| Ludvigsson, 2021 [75]<br>Sweden<br>Prospective cohort<br>Non-industry<br>1 Febrary to 31 July 2020<br>(to 31 July 2020 or death) | People with COVID-19<br>Hospitalized with COVID-19<br>N=365,202 (679 hospitalized)<br>52.7 (16.9)<br>51% | RT-PCR (based on ICD codes) | Age, sex, country, calendar period, education, country of birth, cardiovascular disease, diabetes, chronic obstructive pulmonary disease, end-stage renal disease, liver disease with alcohol use disorder, obesity or dyslipidemia, obstructive sleep apnea, cancer, psychiatric disease | Hospitalization<br>Severe disease (ICU admission or death)<br>ICU admission<br>Mortality |
| Lunski, 2020 [76]<br>USA (Louisiana)<br>Retrospective cohort<br>No funding<br>1 March to 30 April 2020<br>(follow-up NR) | People with COVID-19<br>N=5,145<br><65 y: 74.0%<br>≥65 y: 26.0%<br>39..0% | PCR; Electronic health records (Ochsner's Epic) | Age, sex, race, cancer, chronic kidney disease, COPD, coronary artery disease, diabetes, hypertension, obesity, smoking status (as well as vitals and lab values) | Mortality |

| Author, year;<br>Country (setting);<br>Study design;<br>Funding<br>Study period; follow-up | Enrolled cohort;<br>Study sample;<br>Mean age (SD), years [a]<br>Male, proportion | COVID-19 Diagnosis; Data source | $P^2$ROGRESS risk factors, adjusted for in multivariate regression analysis [b] | Outcomes |
|---|---|---|---|---|
| Mallow, 2020 [77]<br>USA (276 hospitals across USA)<br>Retrospective cohort<br>No funding<br>15 March to 30 April 2020 (follow-up NR) | Hospitalized with COVID-19<br>N=21,676<br>64.9 (17.2)<br>52.8% | Lab-confirmed; Electronic health records (ICD codes) | Age, CDC risk factors, sex, insurance type, chronic lung disease, moderate/severe asthma, heart condition, immunocompromised, obesity, diabetes, chronic kidney disease with dialysis, liver disease, hypertension (as well as statin use, DNR status, hospital status & hospital bed size) | Hospital length of stay<br>ICU admission<br>ICU length of stay<br>Mortality (in-hospital) |
| Mancilla-Galindo, 2021 [78]<br>Mexico<br>Retrospective cohort<br>No funding<br>28 February to 30 May 2020 (follow-up NR) | People with COVID-19<br>N=83,779<br>46.3 (15.9)<br>56.6% | RT-PCR; Federal Government of Mexico database | Age, sex, diabetes, chronic obstructive pulmonary disease, immunosuppression, hypertension, obesity, chronic kidney disease (as well as pneumonia) | Mortality |
| Martinez-Portilla, 2021 (a, b) [79, 80]<br>Mexico | Women with COVID-19<br>N=262,749 (10,366 in matched sample)<br>32.9 (7.5)<br>0% | RT-PCR; Mexican National Registry database | Age, language, nationality, health insurance agency, chronic obstructive pulmonary disease, asthma, smoking, hypertension, cardiovascular disease, diabetes, obesity, chronic renal disease and immunosuppression | Mortality<br>Severe disease (pneumonia)<br>ICU admission<br>Mechanical ventilation |
| Mikami, 2020 [81]<br>USA (New York City, Mount Sinai network)<br>Retrospective cohort<br>NR<br>13 March to 17 April 2020 (follow-up NR) | Hospitalized with COVID-19<br>N=3,708 (2,820 analyzed with an outcome)<br>Median age (IQR)<br>Survivors: 62.0 (49.0-73.0)<br>Non-survivors: 76.0 (65.0-85.0)<br>Survivors: 56.0%<br>Non-survivors: 59.1% | RT-PCR; Electronic health records (Mount Sinai Health system, 8 hospitals and >400 ambulatory practices) | Age, sex, race, cigarette use history, hypertension, diabetes, cancer, azithromycin use, BMI (as well as treatments and lab values) | Mortality (in-hospital) |
| Miller, 2020 [82]<br>USA (metropolitan Detroit, southeast & south-central Michigan)<br>Retrospective cohort<br>Non-industry<br>7 March to 30 April 2020; followed for at least 30 days | Hospitalized with COVID-19<br>N=2,316<br>Alive at 30 days: 62.0 (15.9)<br>Deceased at 30 days: 74.7 (13.8)<br>Alive at 30 days: 50.5%<br>Deceased at 30 days: 57.5% | RT-PCR; Electronic health records (Henry Ford Health System) | Age, sex, residence in low-income area, medicaid insurance, race, COPD, congestive heart failure, coronary artery disease, chronic kidney disease, hypertension, obesity, diabetes mellitus, cancer, dementia, peripheral vascular stroke, stroke | Mortality |

| Author, year;<br>Country (setting);<br>Study design;<br>Funding<br>Study period; follow-up | Enrolled cohort;<br>Study sample;<br>Mean age (SD), years [a]<br>Male, proportion | COVID-19 Diagnosis; Data source | $P^2$ROGRESS risk factors, adjusted for in multivariate regression analysis [b] | Outcomes |
|---|---|---|---|---|
| Misra-Hebert, 2020 [83]<br>USA (Ohio & Florida)<br>Retrospective cohort<br>Industry & Non-industry<br>8 March to 9 June 2020 (follow-up NR) | People with COVID-19<br>N=4,904<br>Median age (IQR)<br>Healthcare worker: 40.6 (30.0-54.0)<br>Non-healthcare worker: 54.5 (39.0-69.0)<br>48.5% | Test-positive; Electronic health records (COVID-19 Cleveland Clinic registry) | Age, sex, race, ethnicity, BMI, asthma, diabetes, hypertension, immunosuppressive disease, median income, population per housing unit, smoking history (as well as presenting symptoms, medications and lab values) | Hospitalization<br>ICU admission |
| Molnar, 2020 [84]<br>USA<br>Prospective cohort<br>NR<br>4 March to 8 May 2020; 5 June 2020 | People with a COVID-19 ICU admission<br>N=4,153<br>Median (IQR): 62.0 (52.0-71.0)<br>64.0% | Test-positive; Electronic medical records | Age; gender; race; ethnicity; pre-existing conditions; immunosuppression (solid organ transplantation at baseline); smoking status | Renal replacement therapy (for acute kidney injury)<br>Mechanical ventilation<br>Mortality |
| Moreira, 2021 [85]<br>USA (nationwide)<br>Prospective cohort<br>Non-industry<br>2 March to 16 July 2020; at least 1 month | Children with COVID-19<br>N=27,045 (20,096 hospitalized)<br>30% 0-9 years, 70% 10-19 years<br>48% | Nasopharyngeal/throat swabs or serologic testing; CDC COVID-NET database | Age, sex, race/ethnicity, presence of comorbidities | Hospitalization<br>Mortality (in hospital) |
| Murillo-Zamora, 2021 [86]<br>Mexico (nationwide)<br>Retrospective cohort<br>No funding<br>4 March to 15 August 2020 | Hospitalized with COVID-19<br>N=66,123<br>3.4% 20-29 years, 16.1% 30-34 years, 33.9% 45-59 years, 46.6% 60+ years<br>60.7% | RT-PCR; Epidemiologic surveillance data | Immunosuppression (any cause except type 2 diabetes and chronic kidney disease), age, sex, tobacco use, obesity, asthma, COPD, type 2 diabetes, arterial hypertension, chronic kidney disease (as well as pneumonia at hospital admission) | Mortality |
| Nachtigall, 2020 [87]<br>Germany (nationwide)<br>Retrospective cohort<br>Non-industry<br>12 February to 12 June 2020; longest follow-up was mean 17.21 days | Hospitalized with COVID-19<br>N=1,904<br>Median (IQR): 73.0 (57.0-82.0)<br>51.5% | RT-PCR; Electronic health records (Helios network) | Age, sex, cardiovascular disease, diabetes, lung disease, malignancy | ICU admission<br>Mechanical ventilation<br>Mortality |
| Nair, 2020 [88]<br>USA (Northwell Health Hospitals in New York) | Hospitalized with COVID-19<br>N=1,707<br>62.9 (11.6) | RT-PCR; Electronic health records | Solid organ transplant, race/ethnicity, body mass index, age, sex, glomerular filtration rate, diabetes, hypertension, coronary artery | Mechanical ventilation<br>Acute kidney injury |

| Author, year;<br>Country (setting);<br>Study design;<br>Funding<br>Study period; follow-up | Enrolled cohort;<br>Study sample;<br>Mean age (SD), years [a]<br>Male, proportion | COVID-19 Diagnosis; Data source | $P^2$ROGRESS risk factors, adjusted for in multivariate regression analysis [b] | Outcomes |
|---|---|---|---|---|
| Retrospective cohort<br>Non-industry<br>1 March to 27 April 2020;<br>to 4 June 2020 | 68.7% | | disease, peripheral vascular disease/peripheral artery disease, heart failure, chronic obstructive pulmonary disease | Severe disease (mechanical ventilation or death) |
| Nijman, 2020 [89]<br>Netherlands (Gelderland and North-Brabant)<br>Prospective cohort<br>March to May 2020;<br>follow-up NR | Hospitalized with COVID-19<br>N=1,006<br>Median (IQR): 69 (58-77)<br>63.9% | RT-PCR; Electronic health records | Age, sex, BMI, diabetes, cardiovascular disease (including hypertension), hypertension, pulmonary disease, immunocompromised (hematological malignancy, stem cell or organ transplant, auto-immune disease, HIV/AIDS, and/or use of immunosuppressive medication) (as well as chronic use of anticoagulant or antiplatelet medication, ACE inhibitors/ angiotensin II receptor blockers, chest x-ray, CT scan severity score, symptoms, lab values) | Mortality (in-hospital death or palliative discharge) |
| Ng, 2020 [90]<br>USA (13 hospitals in a large New York Health system)<br>Retrospective cohort<br>NR<br>1 March to 27 April 2020;<br>27 May 2020 | Hospitalized with COVID-19<br>N=10,482<br>Median (IQR): 66.0 (54.0-77.0)<br>59.5% | RT-PCR; Electronic health records | End-stage kidney disease requiring dialysis, age, sex, race/ethnicity, diabetes, hypertension, cardiovascular diseases (coronary artery disease, heart failure, peripheral vascular disease), respiratory diseases (asthma, COPD), chronic liver disease, cancer, BMI (as well as medication and mechanical ventilation) | Hospital length of stay<br>Mechanical ventilation<br>Mortality (in-hospital) |
| Parra-Bracamonte, 2020 (a, b) [91, 92]<br>Mexico (475 monitoring units in nationwide)<br>Retrospective cohort<br>NR<br>13 January to 17 July 2020<br>(follow-up NR) | People with COVID-19<br>N=331,298 (328,922 analyzed)<br>Median (IQR): 44.0 (33.0-56.0)<br>53.8% | RT-PCR; Open data source of Epidemiologic Surveillance Source of Respiratory Viral Diseases | Age, sex, smoking habits, hypertension, obesity, diabetes, cardiopathy, COPD, asthma, immunosuppressed, chronic kidney disease, other complication | Mortality |
| Patel, 2020 [93]<br>England (UK Biobank)<br>Retrospective cohort<br>Non-industry<br>16 March to 14 April 2020<br>(follow-up NR) | General population<br>N=418,794<br>65.8 (NR)<br>45.0% | RT-PCR; UK Biobank study | Age, sex, race, region, coronary artery disease, hypertension, diabetes, heart failure, ischemic stroke, BMI, COPD, prior pneumonia, Alzheimer's disease/dementia, chronic kidney disease, smoking status, statin use, alcohol consumption, Townsend Index, average income | Hospitalization |

| Author, year;<br>Country (setting);<br>Study design;<br>Funding<br>Study period; follow-up | Enrolled cohort;<br>Study sample;<br>Mean age (SD), years [a]<br>Male, proportion | COVID-19 Diagnosis; Data source | P²ROGRESS risk factors, adjusted for in multivariate regression analysis [b] | Outcomes |
|---|---|---|---|---|
| Petermann-Rocha, 2020 [94]<br>England<br>Prospective cohort<br>Non-industry<br>16 March to 28 June 2020; 28 June 2020 | General population<br>N=383,845<br>67.3 (8.1)<br>55.1% | RT-PCR; UK Biobank, Hospital Episode Statistics and national mortality registers (includes self-report of risk factors) | Age, sex, deprivation index, ethnicity, frailty | Hospitalization |
| Petrilli, 2020 [95]<br>USA (New York City & Long Island)<br>Prospective cohort<br>Non-industry<br>1 March to 8 April 2020; 5 May 2020 | People with COVID-19<br>Hospitalized with COVID-19<br>N=5,279 (2,741 hospitalized)<br>Median (IQR): 54.0 (38.0-66.0)<br><br>49.5% | RT-PCR; Electronic health records | Age, sex, race/ethnicity, history of hypertension, hyperlipidemia, coronary artery disease, heart failure, pulmonary disease (COPD or asthma), malignancy (excluding non-metastatic non-melanoma skin cancer), diabetes, obesity | Hospitalization<br>Severe disease (ICU admission, mechanical ventilation, discharge to hospice or mortality composite, among hospitalized)<br>Mortality (in-hospital or at discharge to hospice) |
| Pinto, 2020 [96]<br>Italy (Hospital of Reggio Emilia)<br>Prospective cohort<br>No funding<br>1 February to 3 April 2020; 30 June 2020 | Hospitalized with COVID-19<br>N=1,226<br>71.7 (14.5)<br>59.8% | RT-PCR; Electronic health records | Age, sex, cancer type, time from cancer diagnosis, and smoking | ICU admission<br>Mortality |
| Poletti, 2020 [97]<br>Italy (Lombardy)<br>Prospective cohort<br>Non-industry<br>February to April 2020; 8 June 2020 | People with COVID-19<br>N=2,824<br>Median (IQR): 53.0 (34.0-64.0)<br>43.2% | RT-PCR & IgG testing; Database of contacts of COVID-19 patients; ongoing serological survey; linelist of COVID-19 patients | Age, sex, cardiovascular disease (including hypertension, hypercholesteremia, myocardiopathy, heart failure, ischemic and valve cardiopathy, vasculopathy) (as well as epidemic period of observed outcomes) | Mortality |
| Polverino, 2020 [98]<br>Italy (nationwide)<br>Retrospective cohort<br>Non-industry<br>25 March to 22 April 2020; follow-up within 30 days from data collection | Hospitalized with COVID-19<br>N=3,179<br>Median (IQR): 69.0 (57.0-78.0)<br>68.3% | RT-PCR; Medical charts | Age, sex, number of comorbidities, clustered by hospital site | Mortality (in-hospital) |

| Author, year;<br>Country (setting);<br>Study design;<br>Funding<br>Study period; follow-up | Enrolled cohort;<br>Study sample;<br>Mean age (SD), years [a]<br>Male, proportion | COVID-19 Diagnosis; Data source | $P^2$ROGRESS risk factors, adjusted for in multivariate regression analysis [b] | Outcomes |
|---|---|---|---|---|
| Portoles, 2020 [99]<br>Spain (Puerta de Hierri University Hospital)<br>Prospective cohort<br>Non-industry<br>25 February to 24 April 2020 (follow-up NR) | Hospitalized with COVID-19<br>N=1,603<br>64.2 (15.6)<br>59.6% | RT-PCR or clinical & tomography scan criteria; Electronic health records | Age, sex, any comorbidity, previous chronic kidney disease | Mortality (in-hospital) |
| Poulson, 2020 [100]<br>USA (nationwide)<br>Retrospective cohort<br>Non-industry<br>5 April to 18 May 2020 (follow-up NR) | People with COVID-19<br>N=124,780<br>0-9 y: 0.5%<br>10-19 y: 1.2%<br>20-29 y: 9.8%<br>30-39 y: 13.5%<br>40-49 y: 14.7%<br>50-59 y: 18.9%<br>60-69 y: 17.8%<br>70-79 y: 12.4%<br>≥80 y: 11.0%<br>48.8% | Lab-confirmed (99.0%); CDC Surveillance Review and Response Group | Age, sex, race, comorbidities | Hospitalization<br>ICU admission<br>Mechanical ventilation<br>Mortality |
| Prado-Galbarro, 2020 [101]<br>Mexico<br>Retrospective cohort<br>No funding<br>27 February to 27 April 2020; 27 April 2020 (median survival 33 days) | People with COVID-19<br>N=15,529<br>≤40 y: 37.4%<br>>40 y: 62.6%<br>57.8% | RT-PCR;<br>Mexican Secretary of Health (nationwide COVID-19 data) | Age, sex, indigenous ethnicity, pneumonia, COPD, immunosuppressive diseases, additional comorbidity, cardiovascular disease, chronic diseases interaction (hypertension, diabetes, obesity, diabetes + hypertension, obesity + hypertension, diabetes + obesity, diabetes + obesity + hypertension), chronic kidney disease (as well as ICU region density & mode of transport) | Mortality |
| Preston, 2021 [102]<br>USA (189 medical facilities)<br>Retrospective cohort<br>NR<br>1 March to 31 October 2020 | Hospitalized with COVID-19<br>N=2,430<br>26% 0-1 years, 10% 2-5 years, 11% 6-11 years, 54% 12-18 years<br>44.6% | Unspecified lab test; Premier Healthcare Database Special COVID-19 Release | Age, sex, race, presence of a chronic condition, insurance | Severe disease (ICU admission, mechanical ventilation, death) |
| Price-Haywood, 2020 [103] | People with COVID-19<br>Hospitalized with COVID-19 | RT-PCR; Electronic health records | Age, sex, race, ethnicity, insurance plan, chronic conditions, BMI, residential zip codes | Hospitalization<br>Mortality (in-hospital) |

| Author, year;<br>Country (setting);<br>Study design;<br>Funding<br>Study period; follow-up | Enrolled cohort;<br>Study sample;<br>Mean age (SD), years [a]<br>Male, proportion | COVID-19 Diagnosis; Data source | $P^2$ROGRESS risk factors, adjusted for in multivariate regression analysis [b] | Outcomes |
|---|---|---|---|---|
| USA (Louisiana, Ochsner Health)<br>Retrospective cohort<br>NR<br>1 March to 11 April 2020; 7 May 2020 | N=3,481 (1,382 hospitalized)<br>54.0 (NR)<br>40.0% | | | |
| Rapp, 2020 [104]<br>USA (New York City, Mount Sinai Network)<br>Retrospective cohort<br>Non-industry<br>29 February to 19 May 2020 (follow-up NR) | Hospitalized with COVID-19<br>N=4,062<br><40 y: 6.6%<br>40-69 y: 47.9%<br>≥70 y: 45.5%<br>57.4% | RT-PCR; Electronic health records (Mount Sinai Hospital System) | Age, sex, asthma, BMI, cancer, COPD, chronic kidney disease, coronary artery disease, HIV, hypertension, smoking (as well as clinical presentation at admission) | Mortality |
| Reilev, 2020 [105]<br>Denmark (nationwide)<br>Retrospective cohort<br>NR<br>27 February to 19 May 2020; 30 days follow-up | People with COVID-19<br>N=11,122<br>Median (IQR): 48.0 (33.0-62.0)<br>42.0% | RT-PCR; Danish Microbiology Database and Electronic Health Records | Age, sex, authorized HCW, number of comorbidities, current drug use, medical history, chronic lung disease, hypertension, ischaemic heart disease, heart failure, atrial fibrillation, stroke, diabetes, dementia, any cancer, chronic liver disease, hospital-diagnosed kidney disease, alcohol abuse, substance abuse, major psychiatric disorder, organ transplantation, medical overweight and obesity, rheumatoid arthritis/connective-tissue disease | Hospitalization<br>Mortality |
| Rentsch, 2020 [106]<br>USA<br>Retrospective cohort<br>Non-industry<br>8 February to 21 June 2020; 22 July 2020 | People with COVID-19<br>N=16,317<br>20-39 y: 17.0%<br>40-49 y: 12.0%<br>50-59 y: 19.0%<br>60-69 y: 23.0%<br>89.6% | Lab-confirmed;<br>US Veterans Health Administration database | Age, sex, race/ethnicity, residence (urban vs. rural), asthma, cancer, chronic kidney disease, chronic pulmonary disease, diabetes, hypertension, liver disease, vascular disease, substance use<br>(as well as medication history) | Mortality |
| Rey, 2020 [107]<br>Spain (127 centers)<br>Retrospective cohort<br>NR | People with COVID-19<br>N=3,080<br>62.3 (20.3)<br>54.8% | RT-PCR; Electronic health records | Age, sex, pre-existing conditions | Mortality |

| Author, year;<br>Country (setting);<br>Study design;<br>Funding<br>Study period; follow-up | Enrolled cohort;<br>Study sample;<br>Mean age (SD), years [a]<br>Male, proportion | COVID-19 Diagnosis; Data source | $P^2$ROGRESS risk factors, adjusted for in multivariate regression analysis [b] | Outcomes |
|---|---|---|---|---|
| 1 March to 20 April 2020; at least 30 days from diagnosis | | | | |
| Rios-Silva, 2020 [108]<br>Mexico<br>Retrospective cohort<br>No funding<br>28 February to 25 May 2020; followed up to 60 days for mortality | Women with COVID-19<br>N=18,390<br>Median (IQR): 36.0 (29.0-43.0)<br>0.0% | RT-qPCR; Mexico Ministry of Health surveillance database | Age, pregnancy status, with individual comorbidities: diabetes, COPD, asthma, immunosuppression, hypertension, obesity, chronic kidney disease, cardiovascular disease, smoking, other comorbidity | Mortality |
| Rodilla, 2020 [109]<br>Spain (nationwide)<br>Retrospective cohort<br>No funding<br>1 March to 24 June 2020 (follow-up NR) | Hospitalized with COVID-19<br>N=12,226<br>67.5 (16.1)<br>57.4% | RT-PCR; SEMI-COVID-19 Network | Age, sex, Charlson Comorbidity Index, atrial fibrillation, chronic kidney disease, heart failure, hypertension (as well as prior treatments with ACEIs & ARBs) | Severe disease (invasive/non-invasive ventilation & ICU admission)<br>Mortality (in-hospital) |
| Samuels, 2021 [110]<br>USA<br>Retrospective cohort<br>No funding<br>2 March to 31 May 2020; follow-up NR | People with COVID-19<br>N=1,692<br>51.0 (19.2)<br>47.4% | RT-PCR; electronic medical records (Memorial Health System) | Age, sex, race/ethnicity, smoking status, flu vaccine status, comorbidity score, chronic cardiac disease, hypertension, obesity, diabetes, kidney disease, rheumatologic disease, hypothyroidism, dementia, malignant neoplasm, hematologic disease, immunosuppressants (as well as ACE inhibitors, ARBs, spironolactone, immunosuppressant, history of being seen in the past 7 days for symptoms, respiratory rate > 24 breaths/min., temperature ≥ 38.0 °C) | ICU admission |
| Sapey, 2020 [111]<br>United Kingdom (University Hospitals Birmingham NHS Foundation Trust)<br>Retrospective cohort<br>Non-industry<br>10 March to 17 April 2020; 12 May 2020 | Hospitalized with COVID-19<br>N=2,217<br>Median (IQR): 73.0 (58.0-84.0)<br>58.2% | RT-PCR; Electronic health records | Age, sex, race, deprivation index, number of comorbidities | Mortality |

| Author, year;<br>Country (setting);<br>Study design;<br>Funding<br>Study period; follow-up | Enrolled cohort;<br>Study sample;<br>Mean age (SD), years [a]<br>Male, proportion | COVID-19 Diagnosis; Data source | P²ROGRESS risk factors, adjusted for in multivariate regression analysis [b] | Outcomes |
|---|---|---|---|---|
| Shah, 2020 (a) [112]<br>United Kingdom<br>Retrospective cohort<br>NR<br>13 March to 15 April 2020; 20 May 2020 | Hospitalized with COVID-19<br>N=1,183<br>Median (IQR): 71.0 (56.0-82.0)<br>57.7% | RT-PCR; Electronic health records (King's College Hospital NHS Foundation Trust) | Age, sex | Mortality |
| Shah, 2020 (b) [113]<br>Scotland<br>Nested case-control<br>Non-industry<br>1 March to 6 June 2020; followed for 28 days for mortality | General population<br>N=3,186,184 (181,375 HCW vs household member)<br>Healthcare worker: 44.5 (11.6)<br>Household member: 30.9 (20.9)<br>Healthcare worker: 21.3%<br>Household member: 61.6% | Lab test positive; Scottish Workforce Information Standard System (SWISS); General Practitioner Contractor Database (GPCD); Community Health Index (CHI); REACT-COVID-19 | Age, sex, socioeconomic deprivation, ethnicity, comorbidities, occupation, part-time status | Hospitalization |
| Tartof, 2020 [114]<br>USA (California)<br>Retrospective cohort<br>Industry<br>13 Febraury to 2 May 2020; followed for 21 days for mortality | People with COVID-19<br>N=6,916<br>49.1 (16.6)<br>45.0% | Lab-confirmed (82% PCR) or diagnostic codes; Kaiser Permanente Southern California health records | Age, sex, race/ethnicity, comorbidities, substance use, neighborhood-level factors, lab value related to diabetes status, (as well as prior medication and health care use, and time) | Mortality |
| Ungaro, 2021 [115]<br>USA (Mount Sinai Hospital System, New York)<br>Retrospective cohort<br>Non-industry<br>1 March to 12 May 2020; follow-up NR | People with COVID-19<br>N=6,792<br>Median (IQR): 63.0 (51.0-73.0) in inflammatory disease; 62.0 (49.0-74.0) in controls<br>54.6% | RT-PCR; Electronic health records | Autoimmune and chronic inflammatory disease (autoimmune hepatitis, ankylosing spondylitis, Sjögren syndrome, scleroderma, psoriasis/psoriatic arthritis, systemic lupus erythematosus, rheumatic arthritis, inflammatory bowel disease, systemic vasculitis, myositis), age, sex, race, comorbidities, obesity | Mechanical ventilation<br>Severe disease (mechanical ventilation or death)<br>Mortality |
| Valenzuela, 2020 [116]<br>USA (Long Island, New York)<br>Retrospective cohort<br>NR | People with COVID-19<br>Hospitalized with COVID-19<br>N=2,039 (996 hospitalized)<br>Median (IQR): 52.0 (38.0-65.0)<br>53.0% | RT-PCR; Electronic health records | Age, sex, race, clinical symptoms, number of comorbidities, and insurance | Hospitalization<br>ICU admission (among hospitalized)<br>Mechanical ventilation (among hospitalized)<br>Mortality (among hospitalized) |

| Author, year;<br>Country (setting);<br>Study design;<br>Funding<br>Study period; follow-up | Enrolled cohort;<br>Study sample;<br>Mean age (SD), years [a]<br>Male, proportion | COVID-19 Diagnosis; Data source | $P^2$ROGRESS risk factors, adjusted for in multivariate regression analysis [b] | Outcomes |
|---|---|---|---|---|
| 7 March to 23 May 2020 (follow-up NR) | | | | |
| van Gerwen, 2020 (a, b) [117, 118]<br>USA (New York City)<br>Retrospective cohort<br>No funding<br>1 March to 1 April 2020; 13 May 2020 | People with COVID-19<br>Hospitalized with COVID-19<br>N=3,703 (2,015 hospitalized)<br>56.8 (18.2)<br>55.3% | RT-PCR; Electronic health records | Age, gender, race, BMI, smoking status, and number of comorbidities | Hospitalization<br>Mechanical ventilation (among hospitalized)<br>Mortality (among hospitalized)<br>Severe disease (mechanical ventilation or mortality) |
| Wang, 2020 [119]<br>USA (New York City)<br>Retrospective cohort<br>No funding<br>24 February to 15 April 2020; 15 April 2020 | People with a COVID-19 hospitalization<br>N=3,273<br>Median (IQR): 65.0 (53.0-77.0)<br>57.3% | RT-PCR & small number by clinical or exposure history;<br>Electronic medical records (Mount Sinai Health System; variables based on self-report, diagnosis code, clinic visit) | Age, sex, race, BMI, smoking status, asthma, COPD, hypertension, obesity, diabetes, HIV, cancer (as well as ICU admission, lab tests duration of stay & vitals) | Mortality (in-hospital) |
| Yehia, 2020 [120]<br>USA<br>Retrospective cohort<br>NR<br>19 February to 31 May 2020; 25 June 2020 | Hospitalized with COVID-19<br>N=7,139<br>Median (IQR): 68.0 (56.0-79.0)<br>51.3% | PCR; Electronic health records | Age, sex, race, obesity, asthma, chronic kidney disease, COPD, congestive heart failure, coronary artery disease, diabetes, Neighborhood Deprivation Index, insurance | Mortality (in-hospital) |
| Zafari, 2020 [121]<br>Iran (studies from any country included)<br>Systematic review | β-Thalassemia patients with COVID-19<br>N=34<br>Range 21-66<br>32% | NR; case series | β-Thalassemia | Hospitalization<br>Mechanical ventilation<br>Mortality |
| Zambrano, 2020 [122]<br>USA<br>Prospective cohort<br>NR<br>22 January to 3 October 2020; 28 October 2020 | Women with COVID-19<br>N=409,462<br>15-24 y: 34.1%<br>25-34 y: 35.4%<br>35-44 y: 30.6%<br>0.0% | Lab-confirmed or antigen test; CDC reports through national COVID-19 case surveillance or National Notifiable Disease Surveillance System | Age, race/ ethnicity, and pre-existing conditions, obesity | ICU admission<br>Mechanical ventilation<br>Mortality |

| Author, year;<br>Country (setting);<br>Study design;<br>Funding<br>Study period; follow-up | Enrolled cohort;<br>Study sample;<br>Mean age (SD), years [a]<br>Male, proportion | COVID-19 Diagnosis; Data source | $P^2$ROGRESS risk factors, adjusted for in multivariate regression analysis [b] | Outcomes |
|---|---|---|---|---|
| Zhu, 2020 [123]<br>United Kingdom<br>Retrospective cohort<br>Non-industry<br>16 March to 16 April 2020 (follow-up NR) | People with COVID-19<br>N=489,769<br>Median (IQR): 58.0 (50.0-63.0)<br>45.0% | PCR; UK Biobank data | Age, sex, race/ethnicity, cardiovascular disease, diabetes and hypertension, household income | Hospitalization |

[a] Values for age are mean (SD), unless otherwise specified.

[b] Risk factors adjusted for in multivariate analysis may differ for outcome(s) reported within a study.

Abbreviations: BMI=body mass index (kg/$m^2$); COPD=chronic obstructive pulmonary disease; HDU=high-dependency unit; HIV=human immunodeficiency virus; ICU=intensive care unit; IQR=interquartile range; NR=not reported; qSOFA=quick sequential organ failure assessment; RT-PCR=reverse transcriptase polymerase chain reaction; SD=standard deviation; y=year(s)

**B. Characteristics of Canadian reports, n=11**

| Author year<br>Design (setting, time period)<br>Funding source | Participants used in analysis | Risk factors and outcomes | Analysis details |
|---|---|---|---|
| **Fisman 2020a** [124]<br>Retrospective cohort (Ontario, Jan-Mar 2020)<br>Funding source: CIHR | 1,734 individuals (43% male, median age 55 y, IQR 40 y) from the general population diagnosed with SARS-CoV-2 via RT-PCR. Data source: iPHIS data system. | Risk factors: age (per 10 year increment); sex; income; long-term care residency; healthcare worker; homeless shelter worker; homelessness; smoking; pregnancy status; anemia or hemoglobinopathy; chronic liver disease; renal disease; diabetes; COPD; asthma; CVD; malignancy; immune compromised; tuberculosis; obesity (not all adjusted for in analysis).<br><br>Outcome: mortality in those with COVID-19 | Multivariate logistic regression |
| **Fisman 2020b** [125]<br>Epidemiological cohort (Ontario, Mar-Apr 2020)<br>Funding source: CIHR | 79,498 residents of long-term care facilities (sex NR, ~93% aged 65+ y) and 1,731,315 community-living older adults (sex NR, aged >69 y). Data source: Ontario Ministry of Health and Long-term Care; Statistics Canada. | Risk factors: long term care residence<br><br>Outcome: mortality in population | Incidence rate ratio, denominator for long-term care based on facility beds; stratified by age in community-dwelling population. |
| **Indigenous Services Canada 2021** [126]<br>Case series (nationwide, Jan 2021)<br>Funding source: Government of Canada | 9,716 people living on First Nations reserves (sex and age NR) who tested positive for SARS-CoV-2 (diagnostic test NR). Data source: Indigenous Services Canada. | Risk factor: place of residence<br><br>Outcomes: hospitalization & mortality in those with COVID-19 | Descriptive statistics |
| **Liu 2020** [127]<br>Retrospective cohort (Ontario and British Columbia, Jan-Sept 2020)<br>Funding source: International Credential Evaluation Service | 6,431 residents of long term care facilities (33-36% male, mean age 83-84 y) who tested positive for SARS-CoV-2 (diagnostic test NR). Data sources: Public Health Ontario; BC Centre for Disease Control. | Risk factor: long-term care residence<br><br>Outcome: mortality in those with COVID-19 | Descriptive statistics |
| **Money 2021** [128]<br>Prospective cohort (British Columbia, Alberta, Ontario, | 1,839 pregnant and 136,062 non-pregnant women with COVID-19 (RT-PCR). Data source: CAN-COVID Preg database | Risk factor: pregnancy<br><br>Outcome: hospitalization, ICU admission | Unadjusted risk ratio |

| Author year<br>Design (setting, time period)<br>Funding source | Participants used in analysis | Risk factors and outcomes | Analysis details |
|---|---|---|---|
| Quebec, Manitoba, March-Dec 2020)<br>Funding sources: PHAC, CIHR, Better Outcomes Registry & Network Ontario, BC Women's Health Foundation | | | |
| **O'Brien 2020a** [129]<br>Retrospective cohort (nationwide, Jan-Jul 2020)<br>Funding sources: CIHR, NSERC, York University | 100,738 individuals (44% male, age NR) from the general population diagnosed with laboratory-confirmed SARS-CoV-2 (diagnostic test NR) (n=69,409 for mortality, using closed cases). Data source: Statistics Canada COVID-19 dataset; 2016b census to normalize healthcare workforce demographics. | Risk factors: age; sex; healthcare worker<br><br>Outcomes: hospitalization, ICU admission & mortality in those with COVID-19 | Descriptive statistics, stratified by age and sex |
| **O'Brien 2020b** (Statistics Canada) [130]<br>Case series (nationwide, Mar-Jul 2020)<br>Funding source: Statistics Canada | Individuals (sample size, sex, and age NR; 94% ≥65 years) who died from probable or laboratory-confirmed (86%) SARS-CoV-2 (diagnostic test NR). Data sources: Canadian Vital Statistics Death Database. | Risk factors: age; long-term care residence; pre-existing conditions<br><br>Outcome: prevalence of conditions among COVID-19 deaths | Descriptive statistics, stratified by age and sex. |
| **Panetta 2020** [131]<br>Retrospective cohort (Montreal, Quebec, Feb-May 2020)<br>Funding source: Réseau SIDA maladies infectueuses grant; Fonds de recherche santé grant. | 27 infants <1 year (median 89, range 34-193 days; 56% male) with laboratory-confirmed SARS-CoV-2 at a single Hospital (Centre hospitalier universitaire Sainte-Justine) | Risk factors: age; sex; comorbid conditions<br><br>Outcomes: hospitalization, mechanical ventilation, ICU admission | Descriptive characteristics |
| **PHAC COVID-19 Surveillance and Epidemiology Team 2020** [132]<br>Retrospective cohort (nationwide, Jan-Jul 2020)<br>Funding source: PHAC | 106,804 individuals (sex and age NR) from the general population diagnosed with probable or laboratory-confirmed SARS-CoV-2 (diagnostic test NR). Data sources: PHAC; datasets provided by provinces and territories. | Risk factors: age; sex; number of comorbidities (n=6,350); long-term care residence or living in seniors' homes (n=10,150) | Descriptive statistics, stratified by age and/or sex |

| Author year<br>Design (setting, time period)<br>Funding source | Participants used in analysis | Risk factors and outcomes | Analysis details |
|---|---|---|---|
| | | Outcome: age-standardized mortality in population (for geography), mortality in those with COVID-19 (other risk factors) | |
| **Subedi 2020** (Statistics Canada) [133]<br>Epidemiological cohort (nationwide, Mar-Jul 2020)<br>Funding source: Statistics Canada | Canadian population. Data sources: Canadian Vital Statistics Death Database and 2016 Census. | Risk factors: visible minorities (by neighborhood <1%, 1-<10%, 10-<25%, ≥25%); some specific ethnic groups<br><br>Outcome: age-adjusted mortality in population | Descriptive statistics, stratified by age |
| **Wang 2020** [134]<br>Prospective cohort (Montreal and Toronto, Jan-May 2020)<br>Funding source: non-industry | 16,490 individuals (45% male, 51% aged <50 y, 21% aged 50-59 y, 14% aged 60-69 y) from the general population (including long term care residents) diagnosed with laboratory-confirmed SARS-CoV-2 (diagnostic test NR). Data sources: iPHIS | Risk factors: age; sex; long term care residency; living in shelters<br><br>Outcome: mortality in those with COVID-19 | Quasi-Poisson regression for residency, adjusting for age and sex; also restricted analysis to ≥60 y |

BC=British Columbia; CIHR=Canadian Institutes of Health Research; COPD=chronic obstructive pulmonary disease; CVD=cardiovascular disease; ICU=intensive care unit; iPHIS= Public Health Information System in Ontario; IQR=interquartile range; NR=not reported; NSERC=National Sciences and Engineering Research Council; RT-PCR=reverse transcription-polymerase chain reaction; SARS-CoV-2=severe acute respiratory syndrome coronavirus 2; y=years.
